# Effectiveness of COVID-19 Vaccines at Preventing Emergency Department or Urgent Care Encounters and Hospitalizations Among Immunocompromised Adults: An Observational Study of Real-World Data Across 10 US States from August-December 2021

**DOI:** 10.1101/2022.10.20.22281327

**Authors:** Peter J. Embi, Matthew E. Levy, Palak Patel, Malini B. DeSilva, Manjusha Gaglani, Kristin Dascomb, Margaret M. Dunne, Nicola P. Klein, Toan C. Ong, Shaun J. Grannis, Karthik Natarajan, Duck-Hye Yang, Edward Stenehjem, Ousseny Zerbo, Charlene McEvoy, Suchitra Rao, Mark G. Thompson, Deepika Konatham, Stephanie A. Irving, Brian E. Dixon, Jungmi Han, Kristin E. Schrader, Nancy Grisel, Ned Lewis, Anupam B. Kharbanda, Michelle A. Barron, Sue Reynolds, I-Chia Liao, William F. Fadel, Elizabeth A. Rowley, Julie Arndorfer, Kristin Goddard, Kempapura Murthy, Nimish R. Valvi, Zachary A. Weber, Bruce Fireman, Sarah E. Reese, Sarah W. Ball, Allison L. Naleway

**Author notes:** **Corresponding author:** Peter J. Embi, MD, MS, FACP, FACMI, FIAHSI, Vanderbilt University Medical Center, 2525 West End Avenue, Suite 1475, Nashville, TN 37203. **Alternate corresponding author:** Matthew E. Levy, PhD, Public Health & Epidemiology Practice, Westat, 1600 Research Blvd, Rockville, MD 20850. **Disclaimers:** The findings and conclusions in this report are those of the authors and do not necessarily represent the views of the US Centers for Disease Control and Prevention.

## Abstract

**Background:** Immunocompromised (IC) persons are at increased risk for severe COVID-19 outcomes and are less protected by 1-2 COVID-19 vaccine doses than are immunocompetent (non-IC) persons. We compared vaccine effectiveness (VE) against medically attended COVID-19 of 2-3 mRNA and 1-2 viral-vector vaccine doses between IC and non-IC adults.

**Methods:** Using a test-negative design among eight VISION Network sites, VE against laboratory-confirmed COVID-19–associated emergency department (ED) or urgent care (UC) events and hospitalizations from 26 August-25 December 2021 was estimated separately among IC and non-IC adults and among specific IC condition subgroups. Vaccination status was defined using number and timing of doses. VE for each status (versus unvaccinated) was adjusted for age, geography, time, prior positive test result, and local SARS-CoV-2 circulation.

**Results:** We analyzed 8,848 ED/UC events and 18,843 hospitalizations among IC patients and 200,071 ED/UC events and 70,882 hospitalizations among non-IC patients. Among IC patients, 3-dose mRNA VE against ED/UC (73% [95% CI: 64-80]) and hospitalization (81% [95% CI: 76-86]) was lower than that among non-IC patients (ED/UC: 94% [95% CI: 93-94]; hospitalization: 96% [95% CI: 95-97]). Similar patterns were observed for viral-vector vaccines. Transplant recipients had lower VE than other IC subgroups.

**Conclusions:** During B.1.617.2 (Delta) variant predominance, IC adults received moderate protection against COVID-19–associated medical events from three mRNA doses, or one viral-vector dose plus a second dose of any product. However, protection was lower in IC versus non-IC patients, especially among transplant recipients, underscoring the need for additional protection among IC adults.

**Key points:** During Delta variant predominance, immunocompromised (IC) adults received moderate protection against COVID-19-associated medical events from three mRNA doses, but IC patients, especially transplant recipients, were less protected than non-IC patients, underscoring the need for additional protection beyond the primary series.

## INTRODUCTION

Immunocompromised (IC) persons, or those with suppressed humoral or cellular immunity resulting from health conditions or medication use, comprise approximately 3% of the U.S. adult population [1, 2]. IC adults are at increased risk for severe COVID-19 outcomes such as hospitalization and death [3-6] and acquire less protection from two COVID-19 mRNA vaccine doses as do immunocompetent (non-IC) persons [7]. Analyses of real-world data have also demonstrated differential vaccine effectiveness (VE) across subgroups of patients with a range of IC conditions [7]. While a third COVID-19 mRNA vaccine dose has been recommended as part of the primary series for IC persons since August 2021 [8], limited evidence exists regarding VE of three mRNA doses for preventing severe outcomes such as COVID-19–associated emergency department (ED) and urgent care (UC) events or hospitalizations among adults with different types of IC conditions [9, 10].

The objective of this study was to estimate, using electronic health record (EHR)-derived data during a period of B.1.617.2 (Delta) variant predominance, the relative VE against laboratory-confirmed COVID-19-associated ED/UC events and hospitalization of 3 versus 2 mRNA vaccine doses and 2 versus 1 viral-vector vaccine dose(s). Estimates also include absolute VE of each vaccinated group compared with unvaccinated status. VE was calculated separately and compared between IC and non-IC adults and among different subgroups with specific types of IC conditions. Determining the real-world effectiveness of 3 mRNA doses and 2 viral-vector doses will inform recommendations for IC patients. They will also provide a basis for future analyses during periods when other variants predominate and when sufficient data have accrued to examine four- and five-dose regimens among IC patients [8].

## METHODS

### Study Setting and Data Source

The VISION Network is a collaboration between the Centers for Disease Control and Prevention (CDC) and eight U.S. health care systems and research centers in 10 states, with integrated medical, laboratory, and vaccination records: Baylor Scott & White Health (Texas), Columbia University Irving Medical Center (New York), HealthPartners Institute (Minnesota, Wisconsin), Intermountain Healthcare (Utah), Kaiser Permanente Northern California, Kaiser Permanente Northwest (Oregon, Washington), Regenstrief Institute (Indiana), and the University of Colorado [7, 11-15].

Within the framework of a test-negative design [16-19], encounters among persons admitted to one of 391 ED/UC facilities and persons hospitalized (>24 hours) at one of 261 hospitals were eligible if patients were aged ≥18 years with ≥1 discharge diagnosis among a previously published list of qualifying COVID-19–like illness (CLI) diagnoses [11]. CLI was defined as a clinical diagnosis (from the International Classification of Diseases [ICD], 9th and 10th Revisions) of acute respiratory illness (e.g., respiratory failure or pneumonia) or signs or symptoms (e.g., cough, fever, dyspnea, vomiting, or diarrhea) that are associated with COVID-19 [20-22]. CLI encounters included ED/UC events and hospitalizations between August 26, 2021 (14 days after the first U.S. recommendation for a third mRNA COVID-19 vaccine dose as part of the primary series for IC individuals [23]) and December 15–25, 2021 (last date during which the Delta variant accounted for >50% of sequenced viruses at each study site [13]). Repeat ED visits and repeat UC clinic visits (within 24 hours) and or hospital readmissions within 30 days post-discharge were combined and analyzed as single medical visits. This study was reviewed and approved by the institutional review board at Westat, Inc.

### Measures

IC status was defined by the presence (presumed IC) or absence (presumed non-IC) of ≥1 discharge diagnosis for immunocompromising conditions, as previously published [7]. Diagnoses across five categories of conditions were derived from previous studies of large hospital-based or administrative databases: 1) solid malignancies, 2) hematologic malignancies, 3) rheumatologic or inflammatory disorders, 4) other intrinsic immune conditions or immunodeficiencies, and 5) organ or stem cell transplants [24-26] (**Supplementary Table 1**). Data on immunosuppressive medications were not available.

Vaccination status was documented in EHRs or state or local immunization registries [11] and was defined as of the medical event index, or the date of specimen collection associated with the most recent SARS-CoV-2 test result before the ED/UC event or hospitalization (or, if testing only occurred after admission, the date of admission). Primary analyses included recipients of Moderna (mRNA-1273) and/or Pfizer-BioNTech (BNT162b2) products. Patients were classified as unvaccinated, 2-dose mRNA-vaccinated (≥14 days earlier), or 3-dose mRNA-vaccinated (≥7 days earlier). Patients with two doses were further subdivided as <150 and ≥150 days earlier. We excluded mRNA vaccine recipients who received 1) only 1 dose, 2) second dose 1–13 days earlier, 3) third dose 1–6 days earlier, or 4) ≥4 doses. Secondary analyses included Janssen (Johnson & Johnson [Ad26.COV2]) vaccine recipients, with patients classified as 1-dose Janssen-vaccinated (≥14 earlier) or 2-dose vaccinated (≥7 days earlier). Due to sample size constraints, any product was used to define second dose (i.e., mRNA or Janssen). Among Janssen recipients, we excluded those who received 1) first dose 1–13 days earlier, 2) second dose (any product) 1–6 days earlier, or 3) ≥3 doses.

SARS-CoV-2 infection was confirmed by molecular assays (e.g., real-time reverse-transcriptase– polymerase-chain-reaction [RT-PCR] assays) obtained within 14 days before to <72 hours after an ED/UC clinic visit or hospital admission. We only included events with an available SARS-CoV-2 molecular test result during this window. To minimize confounding related to recent infection, patients with a positive SARS-CoV-2 result 15–89 days earlier were excluded. Demographics and underlying medical conditions (defined by ICD codes assigned at medical event) were extracted from EHRs.

### Statistical Analysis

A test-negative design was used to assess VE with respect to an ED/UC event or hospitalization associated with laboratory-confirmed SARS-CoV-2 infection and CLI. VE was estimated by comparing odds of a positive SARS-CoV-2 test result between vaccinated and unvaccinated patients using multivariable logistic regression models [7, 11, 13]. VE was adjusted for age (spline), geographic region, calendar time (spline), prior documented positive SARS-CoV-2 test result (none, 90–364 days ago, ≥365 days ago), and local SARS-CoV-2 circulation (7-day moving average of percentage of RT-PCR tests that were SARS-CoV-2–positive within the medical facility’s geographic region; spline). VE was also weighted for patients’ inverse propensity to be vaccinated (if vaccinated) or unvaccinated (if not vaccinated). Generalized boosted regression trees were used to estimate the propensity to be vaccinated based on sociodemographics, underlying medical conditions, known prior SARS-CoV-2 infection (pre-vaccination, if vaccinated), and facility characteristics. Separate weights were calculated for each VE model and were truncated at the 99^th^ percentile of the distribution of weights. Any covariates with an absolute standardized mean or proportion difference (SMD) ≥0.20 after weighting were also included in the model for the respective VE estimate(s) to minimize residual confounding.

VE estimates were stratified by IC status, vaccine product received, age group (18–64, ≥65 years), and race/ethnicity (non-Hispanic white, non-Hispanic Black, Hispanic). Among IC patients, VE was also calculated separately among five mutually exclusive subgroups of patients with each type of immunocompromising diagnosis (patients with multiple types were excluded) and separately among recipients and non-recipients of organ/stem cell transplants (no patients excluded). To directly compare the 3-dose versus 2-dose mRNA vaccine series, relative VE was calculated using a similar methodology with 2 doses serving as the referent group. Effect modification by IC status (IC vs. non-IC) and type of IC condition (transplant recipient vs. non-transplant recipient) was assessed using interaction terms with vaccination status in regression models. To evaluate mRNA product differences, a contrast was used to test differences in regression coefficients between Moderna and Pfizer-BioNTech. P-values <0.05 were considered statistically significant. Analyses were performed using SAS version 9.4 (SAS Institute) or R software, version 4.1.2 (R Foundation for Statistical Computing).

## RESULTS

### Descriptive Characteristics

A total of 208,919 ED/UC medical events and 89,725 hospitalizations were identified during August 26–December 25, 2021, among adults with CLI and SARS-CoV-2 testing who were either unvaccinated or had received 2-3 mRNA vaccine doses. Characteristics of non-IC and IC patients overall and by vaccination status are provided in **Table 1** for ED/UC events and **Table 2** for hospitalizations. The proportions of all ED/UC events and hospitalizations that were among IC persons were 4% and 21%, respectively. Median age was 47 years (interquartile range [IQR]: 31–66) and 65 years (IQR: 48–77) among non-IC patients in ED/UC and hospital settings, respectively, and 66 years (IQR: 54–75) and 68 years (IQR: 58–77) among IC patients in those settings (data not shown). Overall, proportions of ED/UC events among non-Hispanic Black and Hispanic patients were 9% and 15%, respectively, and 11% and 12% for hospitalizations. Overall, a smaller proportion of ED/UC (19%) than hospitalized (64%) patients had a discharge diagnosis for a chronic respiratory condition. Proportions of ED/UC events and hospitalizations among patients with a historical positive SARS-CoV-2 test result ≥90 days earlier was 6– 7%. There were differences by network site, age, and race/ethnicity between 2-/3-doses mRNA-vaccinated and unvaccinated patients. The application of inverse propensity-to-be-vaccinated weighting reduced differences between vaccinated and unvaccinated patients with respect to these and other factors (**Supplementary Tables 2-3**).

**Table 1.**
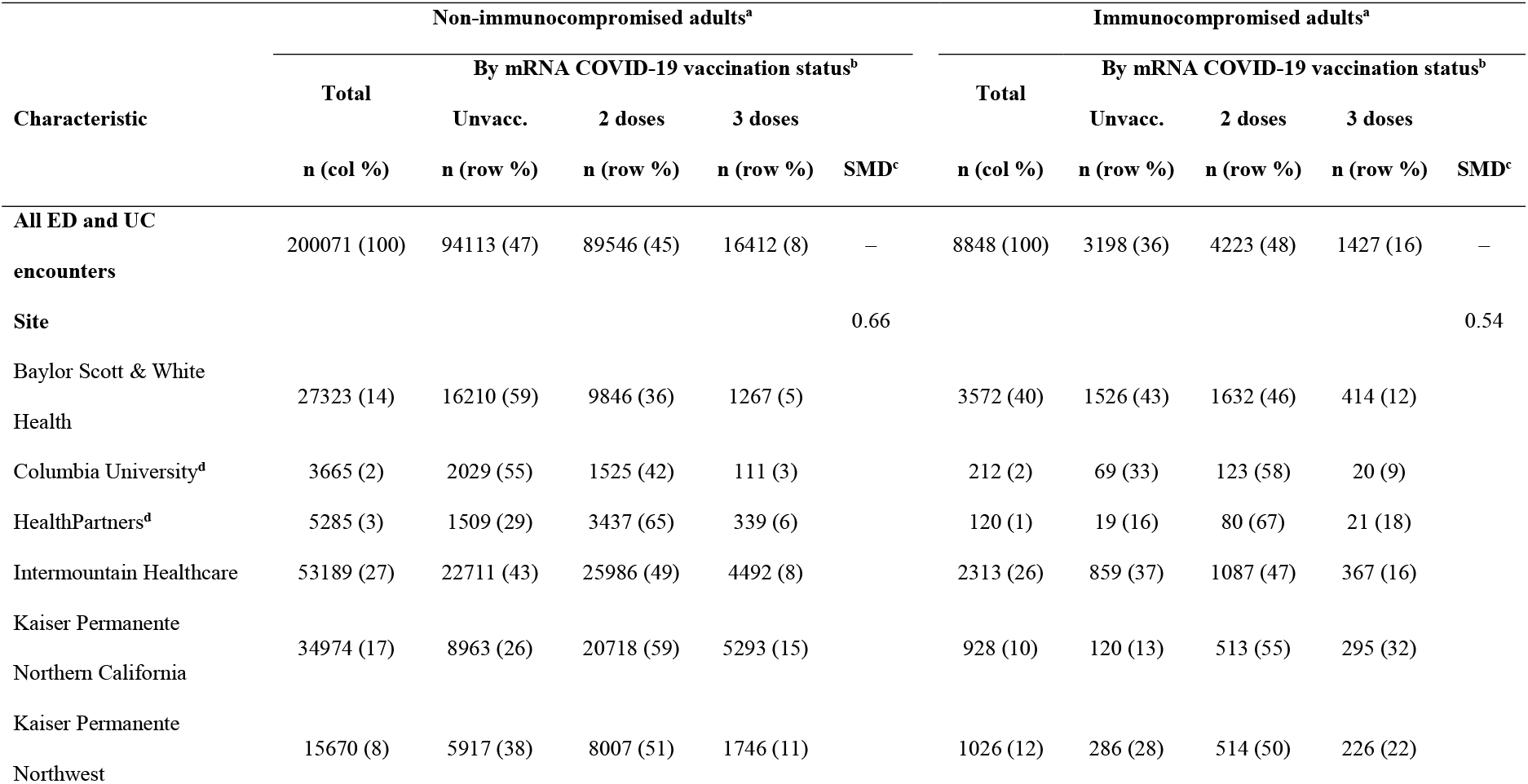
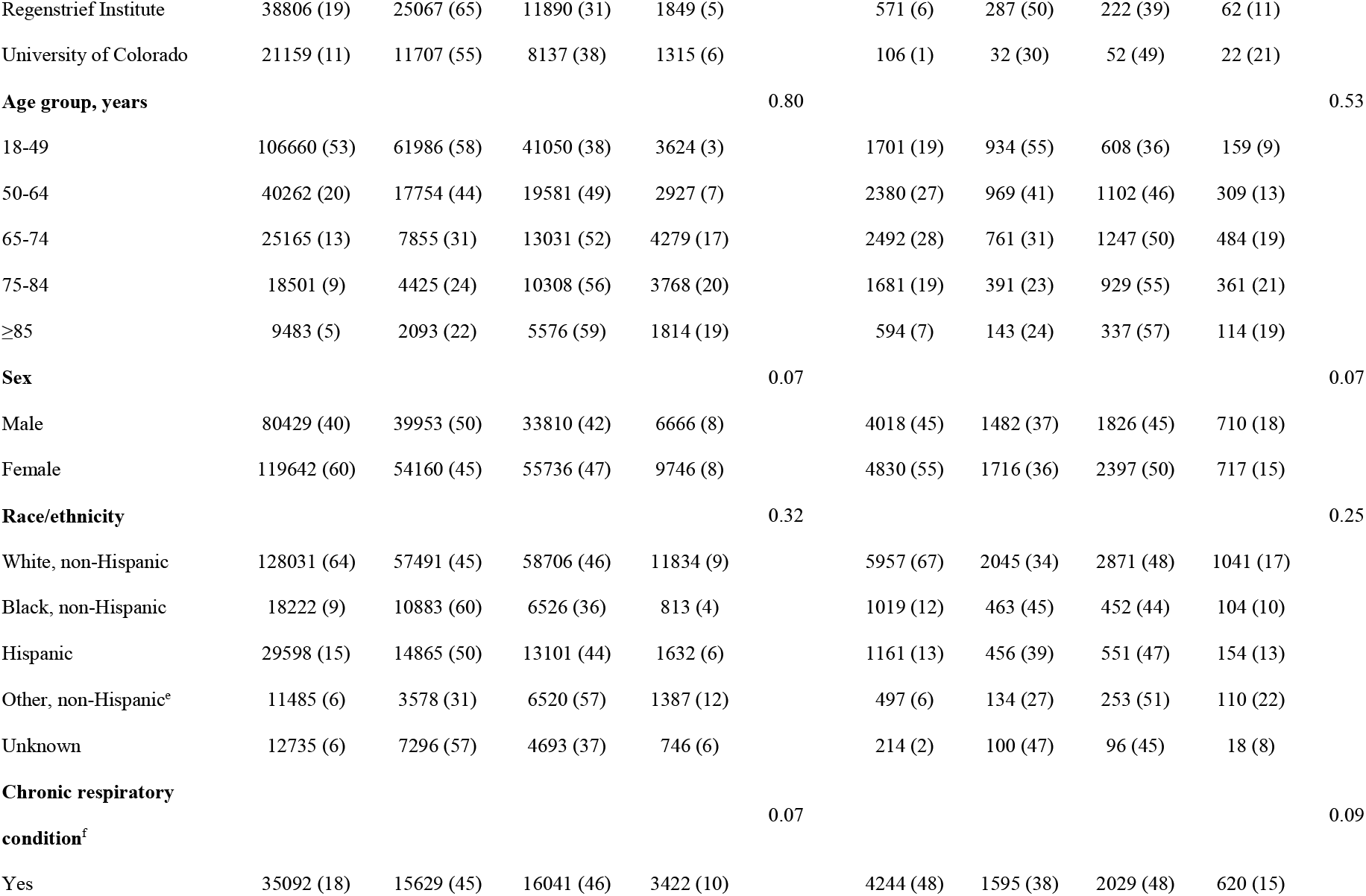
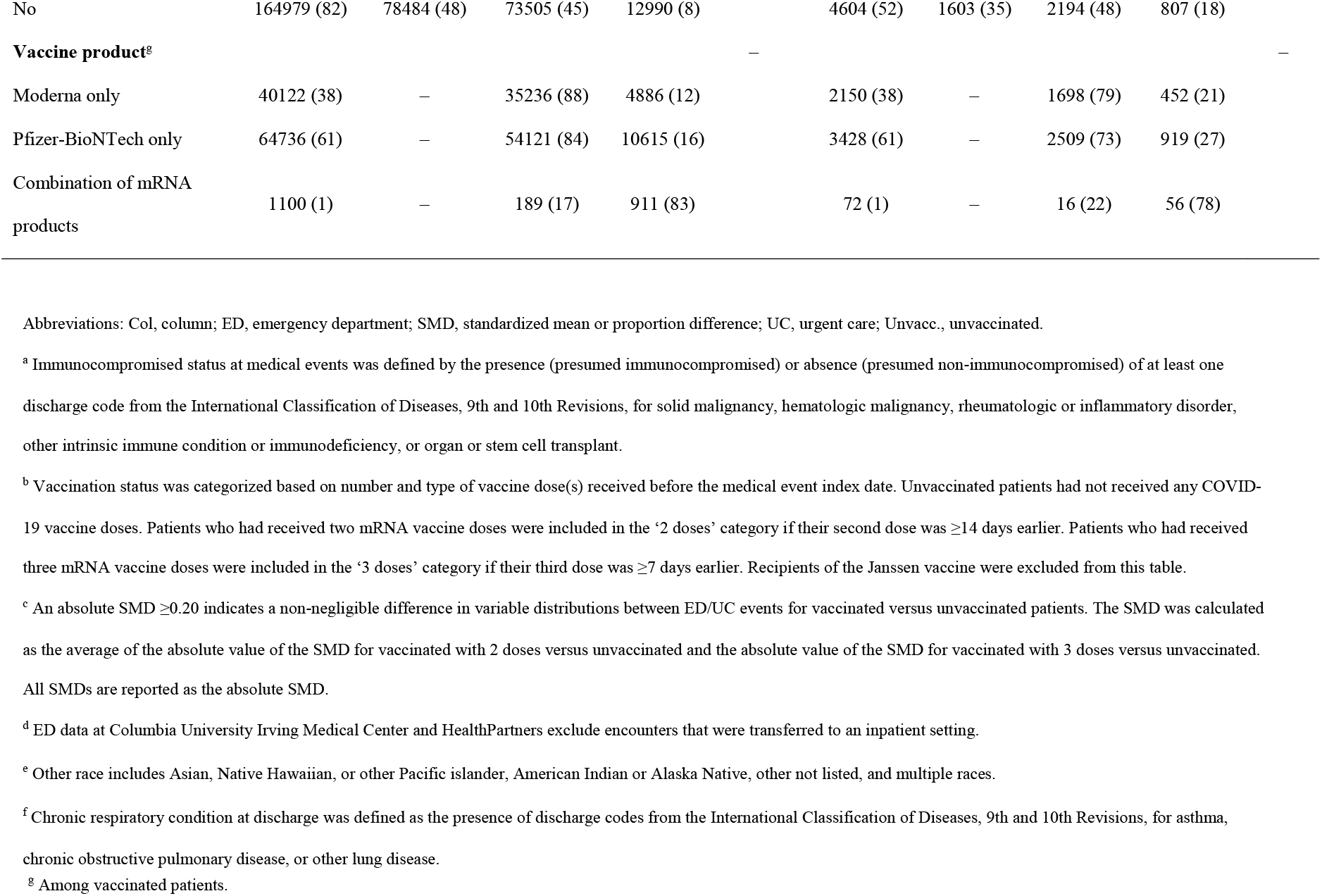
Characteristics of emergency department (ED) and urgent care (UC) events among non-immunocompromised (non-IC) and immunocompromised (IC) adults with COVID-19–like illness, by mRNA COVID-19 vaccination status, VISION Network, 10 states, August– December 2021 (n=208,919).

**Table 2.**
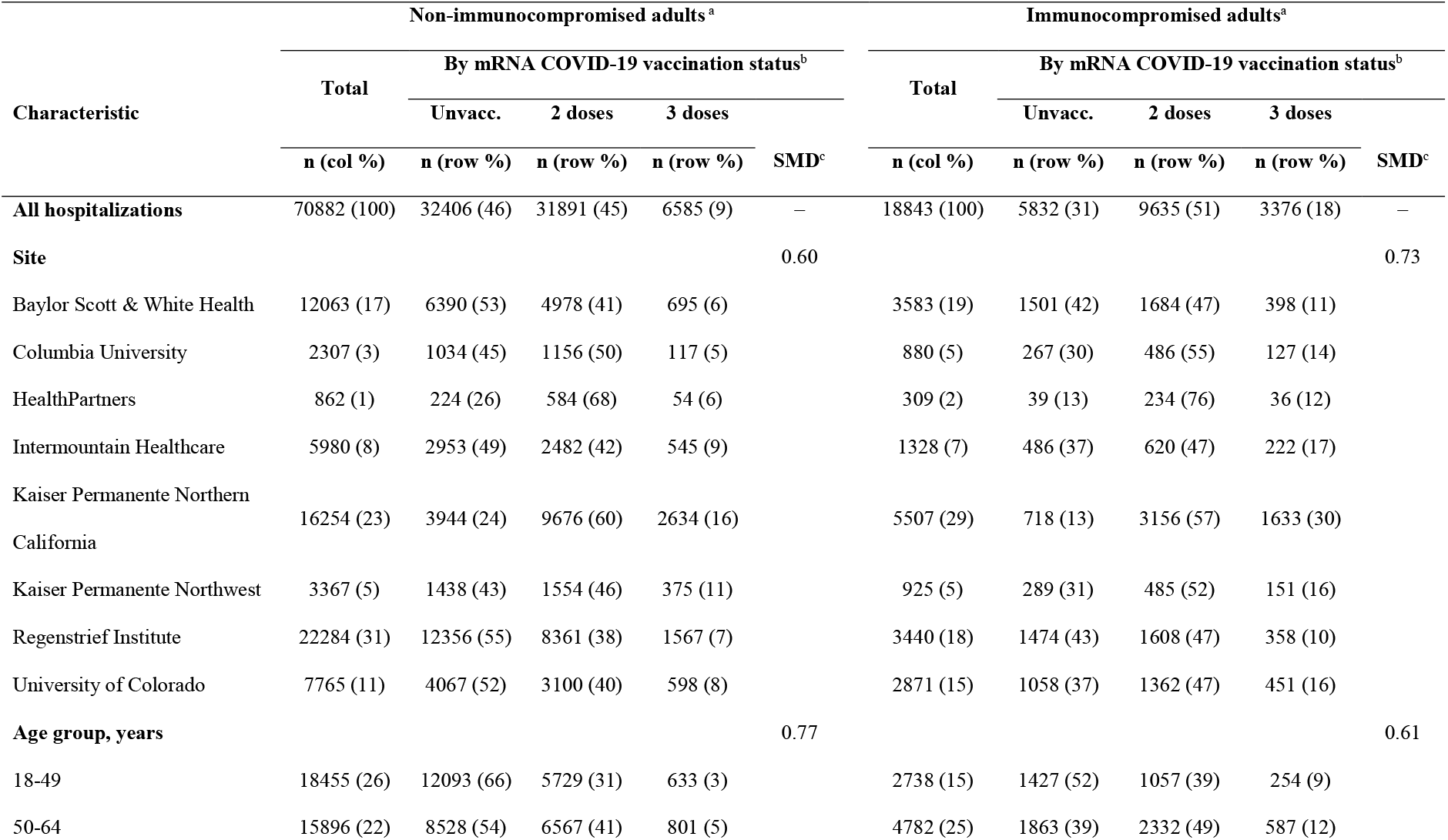
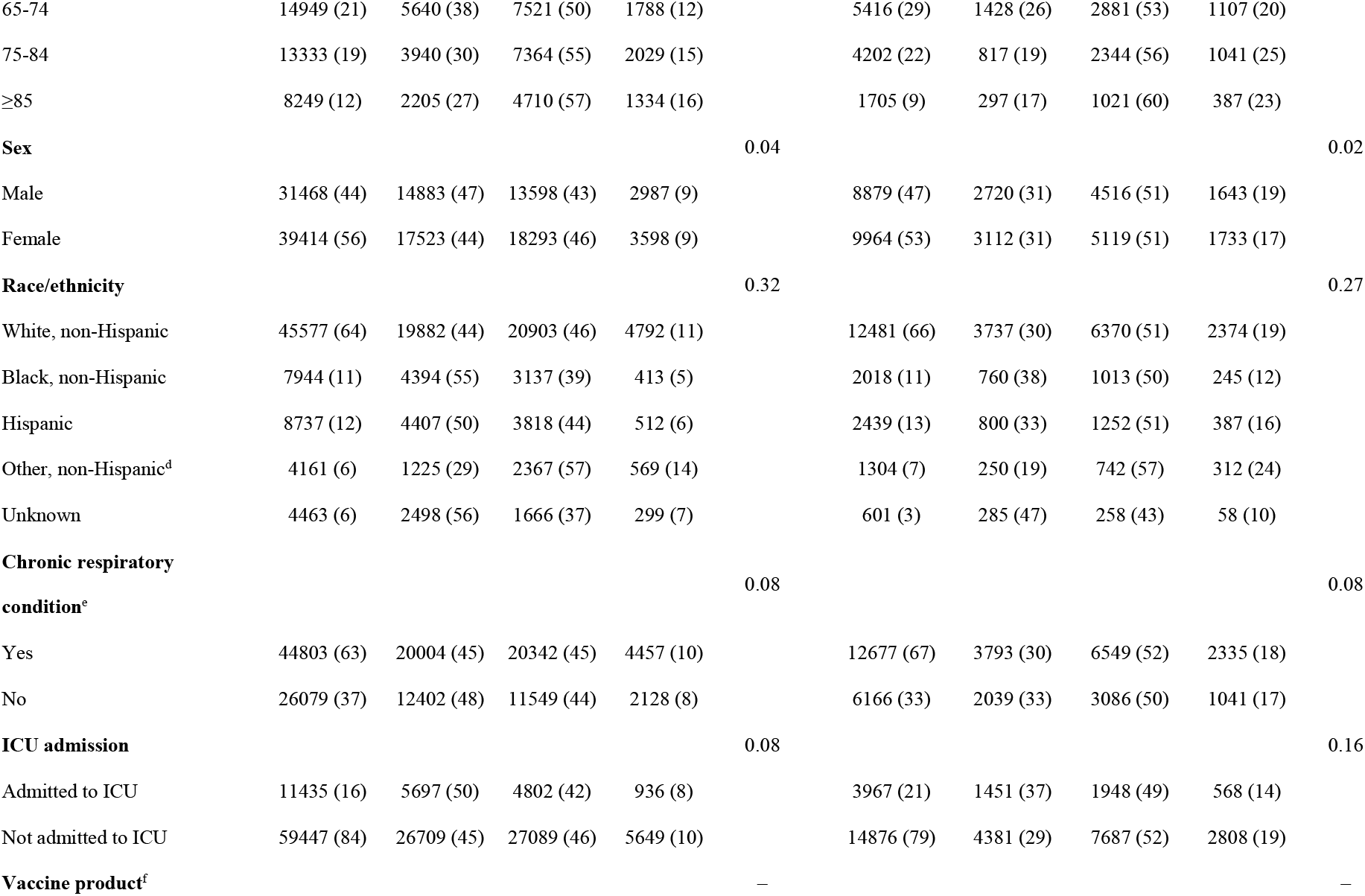
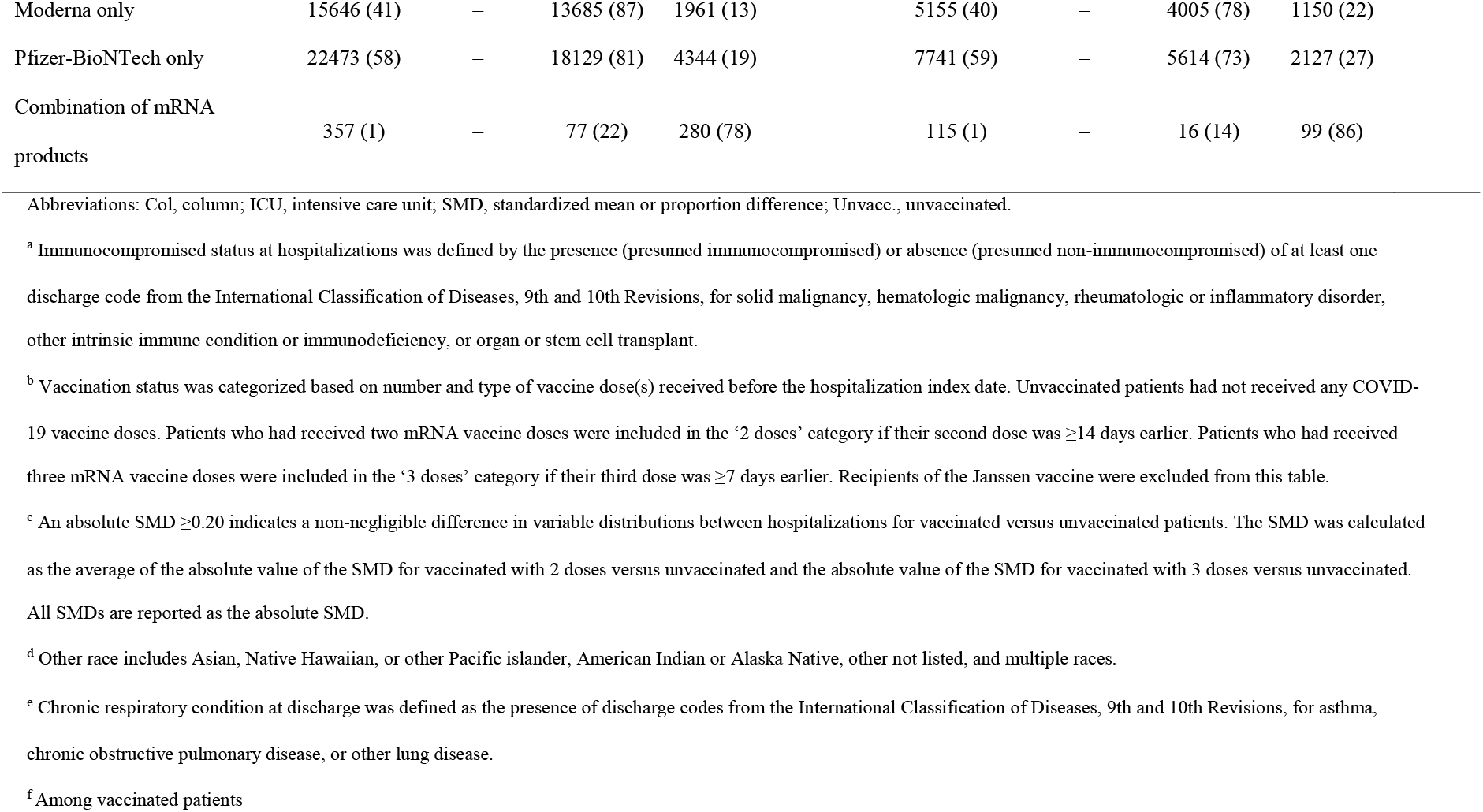
Characteristics of hospitalizations among non-immunocompromised and immunocompromised adults with COVID-19–like illness, by mRNA COVID-19 vaccination status, VISION Network, 10 states, August–December 2021 (n=89,725).

Among 8,848 ED/UC events and 18,843 hospitalizations for IC patients, 36% and 42%, respectively, had a discharge diagnosis for solid malignancy, compared with diagnoses for hematologic malignancy (11%/13%), rheumatologic/inflammatory disorder (27%/25%), another intrinsic immune condition or underlying immunodeficiency (34%/35%), and organ/stem cell transplant (9%/8%) (data not shown). The majority of ED/UC visits (84%) and hospitalizations (79%) had only one of these five types, whereas 14% and 18%, respectively, had exactly two, 1%/2% had three, and <0.1%/0.3% had four or five.

In both settings, a smaller proportion of non-IC patients were vaccinated with two (45% in both settings) or three (8%/9%) mRNA vaccine doses compared with IC patients (two doses: 48%/51%; three doses: 16%/18%) (**Tables 1-2**). Overall, 38% and 41% of vaccinated non-IC patients in ED/UC and hospital settings, received the Moderna vaccine, compared with 61%/58% with the Pfizer-BioNTech vaccine and 1%/1% with a combination of both products. Among vaccinated IC patients, 38% and 40% in ED/UC and hospital settings received the Moderna vaccine, compared with 61%/59% with the Pfizer-BioNTech vaccine and 1%/1% with a combination.

In ED/UC settings, the median time since 2-dose mRNA-vaccinated patients’ second dose was 189 days among non-IC patients (IQR: 146–225) and 196 days among IC patients (IQR: 160–230); median durations in hospitals were 199 days (IQR: 161–233) and 198 days (IQR: 163–231), respectively (data not shown). In ED/UC settings, the median time since 3-dose mRNA-vaccinated patients’ third dose was 35 days among non-IC patients (IQR: 17–49) and 45 days among IC patients (IQR: 26–72); median time in hospitals was 34 days (IQR: 19–54) among non-IC and 43 days (IQR: 24–66) among IC patients. Distributions were similar by mRNA vaccine product.

### mRNA COVID-19 VE by Immunocompromised Status

Among 200,071 ED/UC events for non-IC patients, 21% had laboratory-confirmed SARS-CoV-2 infection (35% infected among unvaccinated; 10% among 2-dose vaccinated; 3% among 3-dose vaccinated), compared with 13% among 8,848 ED/UC events for IC patients (21% infected among unvaccinated; 9% among 2-dose vaccinated; 5% among 3-dose vaccinated). Among IC adults, 2-dose mRNA VE was 67% (95% confidence interval [CI]: 62–72) and 3-dose mRNA VE was 73% (95% CI: 64–80) against SARS-CoV-2 infection at an ED/UC event, compared with 81% (95% CI: 81–82) and 94% (95% CI: 93–94) among non-IC adults (**Table 3**). VE against ED/UC events for 2 and 3 doses was significantly lower among IC versus non-IC patients (tests of interaction, both p<0.05).

**Table 3.**
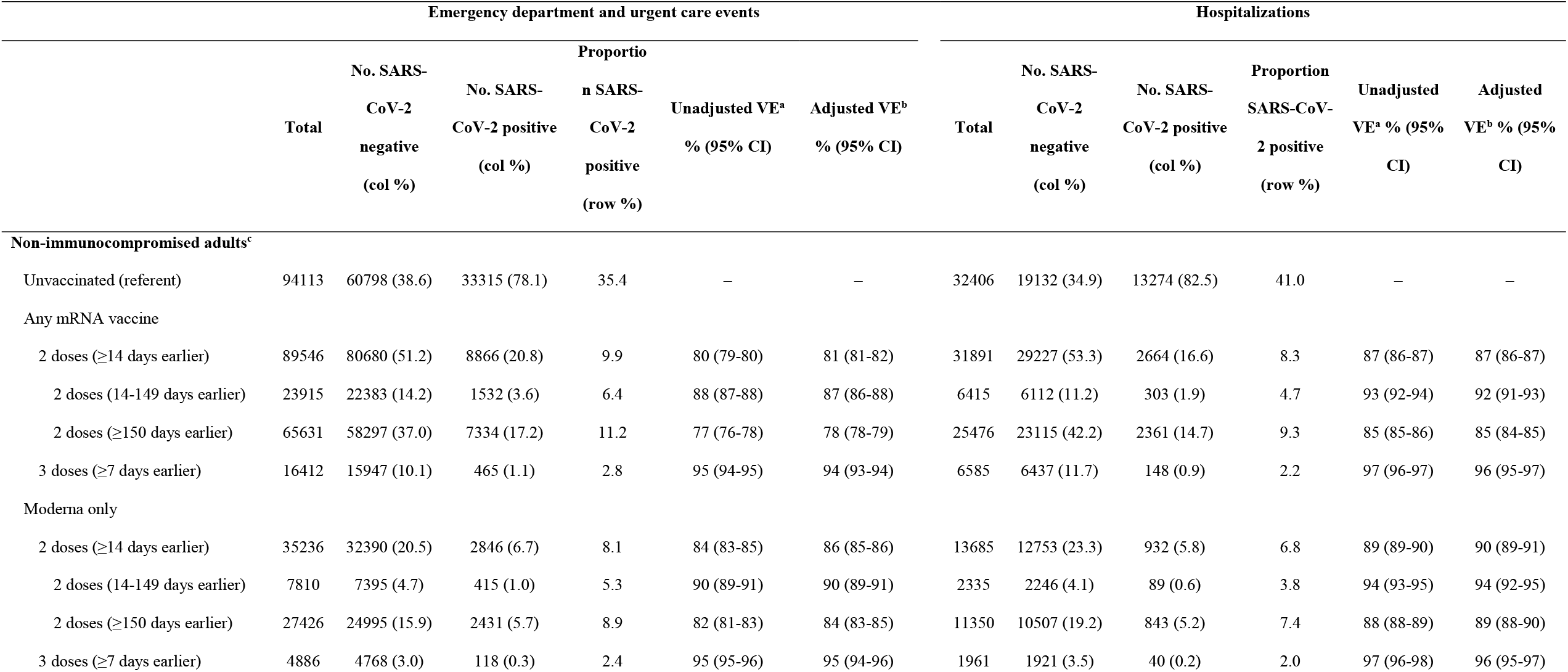
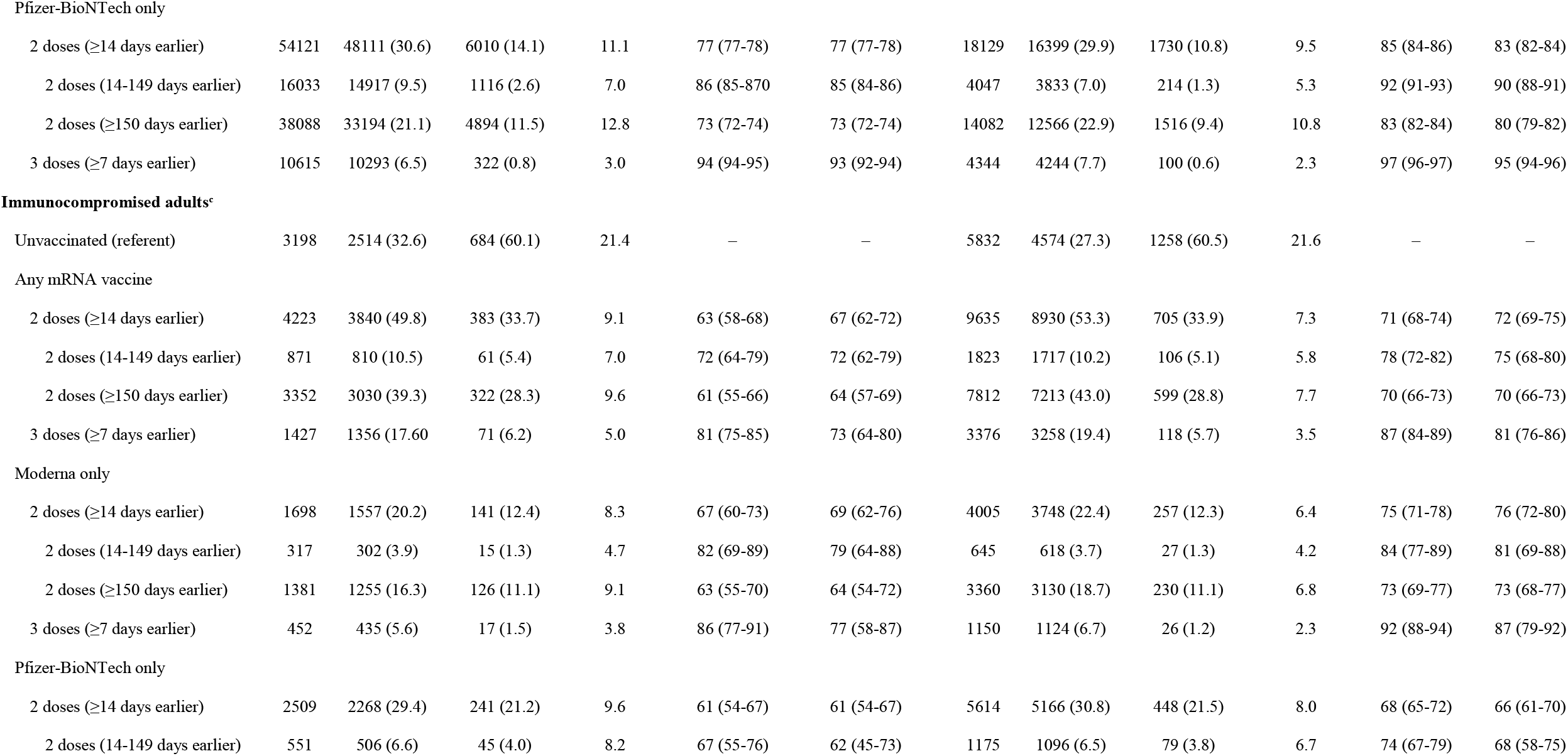
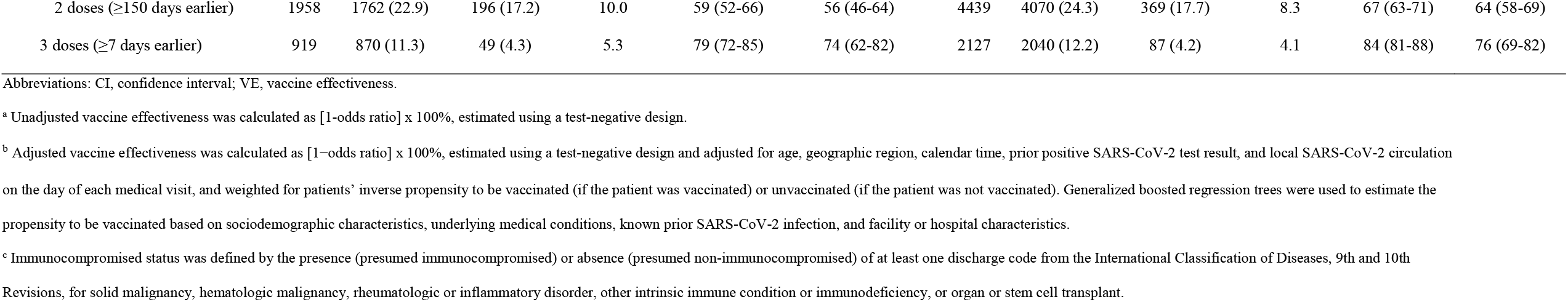
mRNA COVID-19 vaccine effectiveness against laboratory-confirmed COVID-19–associated emergency department or urgent care events and hospitalizations among non-immunocompromised and immunocompromised adults, by number and timing of vaccine doses and vaccine product received, August–December 2021.

| Emergency department and urgent care events |  |  |  |  |  |  | Hospitalizations |  |  |  |  |  |
| --- | --- | --- | --- | --- | --- | --- | --- | --- | --- | --- | --- | --- |
|  | Total | No. SARS-CoV-2 negative (col %) | No. SARS-CoV-2 positive (col %) | Proportion SARS-CoV-2 positive (row %) | Unadjusted VE<sup>a</sup> % (95% CI) | Adjusted VE<sup>b</sup> % (95% CI) | Total | No. SARS-CoV-2 negative (col %) | No. SARS-CoV-2 positive (col %) | Proportion SARS-CoV-2 positive (row %) | Unadjusted VE<sup>a</sup> % (95% CI) | Adjusted VE<sup>b</sup> % (95% CI) |
| Non-immunocompromised adults<sup>c</sup> |  |  |  |  |  |  |  |  |  |  |  |  |
| Unvaccinated (referent) | 94113 | 60798 (38.6) | 33315 (78.1) | 35.4 | — | — | 32406 | 19132 (34.9) | 13274 (82.5) | 41.0 | — | — |
| Any mRNA vaccine |  |  |  |  |  |  |  |  |  |  |  |  |
| 2 doses (≥14 days earlier) | 89546 | 80680 (51.2) | 8866 (20.8) | 9.9 | 80 (79-80) | 81 (81-82) | 31891 | 29227 (53.3) | 2664 (16.6) | 8.3 | 87 (86-87) | 87 (86-87) |
| 2 doses (14-149 days earlier) | 23915 | 22383 (14.2) | 1532 (3.6) | 6.4 | 88 (87-88) | 87 (86-88) | 6415 | 6112 (11.2) | 303 (1.9) | 4.7 | 93 (92-94) | 92 (91-93) |
| 2 doses (≥150 days earlier) | 65631 | 58297 (37.0) | 7334 (17.2) | 11.2 | 77 (76-78) | 78 (78-79) | 25476 | 23115 (42.2) | 2361 (14.7) | 9.3 | 85 (85-86) | 85 (84-85) |
| 3 doses (≥7 days earlier) | 16412 | 15947 (10.1) | 465 (1.1) | 2.8 | 95 (94-95) | 94 (93-94) | 6585 | 6437 (11.7) | 148 (0.9) | 2.2 | 97 (96-97) | 96 (95-97) |
| Moderna only |  |  |  |  |  |  |  |  |  |  |  |  |
| 2 doses (≥14 days earlier) | 35236 | 32390 (20.5) | 2846 (6.7) | 8.1 | 84 (83-85) | 86 (85-86) | 13685 | 12753 (23.3) | 932 (5.8) | 6.8 | 89 (89-90) | 90 (89-91) |
| 2 doses (14-149 days earlier) | 7810 | 7395 (4.7) | 415 (1.0) | 5.3 | 90 (89-91) | 90 (89-91) | 2335 | 2246 (4.1) | 89 (0.6) | 3.8 | 94 (93-95) | 94 (92-95) |
| 2 doses (≥150 days earlier) | 27426 | 24995 (15.9) | 2431 (5.7) | 8.9 | 82 (81-83) | 84 (83-85) | 11350 | 10507 (19.2) | 843 (5.2) | 7.4 | 88 (88-89) | 89 (88-90) |
| 3 doses (≥7 days earlier) | 4886 | 4768 (3.0) | 118 (0.3) | 2.4 | 95 (95-96) | 95 (94-96) | 1961 | 1921 (3.5) | 40 (0.2) | 2.0 | 97 (96-98) | 96 (95-97) |
| Pfizer-BioNTech only |  |  |  |  |  |  |  |  |  |  |  |  |
| 2 doses (≥14 days earlier) | 54121 | 48111 (30.6) | 6010 (14.1) | 11.1 | 77 (77-78) | 77 (77-78) | 18129 | 16399 (29.9) | 1730 (10.8) | 9.5 | 85 (84-86) | 83 (82-84) |
| 2 doses (14-149 days earlier) | 16033 | 14917 (9.5) | 1116 (2.6) | 7.0 | 86 (85-870) | 85 (84-86) | 4047 | 3833 (7.0) | 214 (1.3) | 5.3 | 92 (91-93) | 90 (88-91) |
| 2 doses (≥150 days earlier) | 38088 | 33194 (21.1) | 4894 (11.5) | 12.8 | 73 (72-74) | 73 (72-74) | 14082 | 12566 (22.9) | 1516 (9.4) | 10.8 | 83 (82-84) | 80 (79-82) |
| 3 doses (≥7 days earlier) | 10615 | 10293 (6.5) | 322 (0.8) | 3.0 | 94 (94-95) | 93 (92-94) | 4344 | 4244 (7.7) | 100 (0.6) | 2.3 | 97 (96-97) | 95 (94-96) |
| Unvaccinated (referent) | 3198 | 2514 (32.6) | 684 (60.1) | 21.4 | – | – | 5832 | 4574 (27.3) | 1258 (60.5) | 21.6 | – | – |
| Any mRNA vaccine |  |  |  |  |  |  |  |  |  |  |  |  |
| 2 doses (≥14 days earlier) | 4223 | 3840 (49.8) | 383 (33.7) | 9.1 | 63 (58-68) | 67 (62-72) | 9635 | 8930 (53.3) | 705 (33.9) | 7.3 | 71 (68-74) | 72 (69-75) |
| 2 doses (14-149 days earlier) | 871 | 810 (10.5) | 61 (5.4) | 7.0 | 72 (64-79) | 72 (62-79) | 1823 | 1717 (10.2) | 106 (5.1) | 5.8 | 78 (72-82) | 75 (68-80) |
| 2 doses (≥150 days earlier) | 3352 | 3030 (39.3) | 322 (28.3) | 9.6 | 61 (55-66) | 64 (57-69) | 7812 | 7213 (43.0) | 599 (28.8) | 7.7 | 70 (66-73) | 70 (66-73) |
| 3 doses (≥7 days earlier) | 1427 | 1356 (17.60) | 71 (6.2) | 5.0 | 81 (75-85) | 73 (64-80) | 3376 | 3258 (19.4) | 118 (5.7) | 3.5 | 87 (84-89) | 81 (76-86) |
| Moderna only |  |  |  |  |  |  |  |  |  |  |  |  |
| 2 doses (≥14 days earlier) | 1698 | 1557 (20.2) | 141 (12.4) | 8.3 | 67 (60-73) | 69 (62-76) | 4005 | 3748 (22.4) | 257 (12.3) | 6.4 | 75 (71-78) | 76 (72-80) |
| 2 doses (14-149 days earlier) | 317 | 302 (3.9) | 15 (1.3) | 4.7 | 82 (69-89) | 79 (64-88) | 645 | 618 (3.7) | 27 (1.3) | 4.2 | 84 (77-89) | 81 (69-88) |
| 2 doses (≥150 days earlier) | 1381 | 1255 (16.3) | 126 (11.1) | 9.1 | 63 (55-70) | 64 (54-72) | 3360 | 3130 (18.7) | 230 (11.1) | 6.8 | 73 (69-77) | 73 (68-77) |
| 3 doses (≥7 days earlier) | 452 | 435 (5.6) | 17 (1.5) | 3.8 | 86 (77-91) | 77 (58-87) | 1150 | 1124 (6.7) | 26 (1.2) | 2.3 | 92 (88-94) | 87 (79-92) |
| Pfizer-BioNTech only |  |  |  |  |  |  |  |  |  |  |  |  |
| 2 doses (≥14 days earlier) | 2509 | 2268 (29.4) | 241 (21.2) | 9.6 | 61 (54-67) | 61 (54-67) | 5614 | 5166 (30.8) | 448 (21.5) | 8.0 | 68 (65-72) | 66 (61-70) |
| 2 doses (14-149 days earlier) | 551 | 506 (6.6) | 45 (4.0) | 8.2 | 67 (55-76) | 62 (45-73) | 1175 | 1096 (6.5) | 79 (3.8) | 6.7 | 74 (67-79) | 68 (58-75) |
| 2 doses (≥150 days earlier) | 1958 | 1762 (22.9) | 196 (17.2) | 10.0 | 59 (52-66) | 56 (46-64) | 4439 | 4070 (24.3) | 369 (17.7) | 8.3 | 67 (63-71) | 64 (58-69) |
| 3 doses (≥7 days earlier) | 919 | 870 (11.3) | 49 (4.3) | 5.3 | 79 (72-85) | 74 (62-82) | 2127 | 2040 (12.2) | 87 (4.2) | 4.1 | 84 (81-88) | 76 (69-82) |

Abbreviations: CI, confidence interval; VE, vaccine effectiveness.
<sup>a</sup> Unadjusted vaccine effectiveness was calculated as [1-odds ratio] x 100%, estimated using a test-negative design.
<sup>b</sup> Adjusted vaccine effectiveness was calculated as [1-odds ratio] x 100%, estimated using a test-negative design and adjusted for age, geographic region, calendar time, prior positive SARS-CoV-2 test result, and local SARS-CoV-2 circulation on the day of each medical visit, and weighted for patients’ inverse propensity to be vaccinated (if the patient was vaccinated) or unvaccinated (if the patient was not vaccinated). Generalized boosted regression trees were used to estimate the propensity to be vaccinated based on sociodemographic characteristics, underlying medical conditions, known prior SARS-CoV-2 infection, and facility or hospital characteristics.
<sup>c</sup> Immunocompromised status was defined by the presence (presumed immunocompromised) or absence (presumed non-immunocompromised) of at least one discharge code from the International Classification of Diseases, 9th and 10th Revisions, for solid malignancy, hematologic malignancy, rheumatologic or inflammatory disorder, other intrinsic immune condition or immunodeficiency, or organ or stem cell transplant.

Among 70,882 hospitalizations for non-IC patients, 22% had SARS-CoV-2 infection (41% infected among unvaccinated; 8% among 2-dose vaccinated; 2% among 3-dose vaccinated), compared with 11% among 18,843 hospitalizations for IC patients (22% infected among unvaccinated; 7% among 2-dose vaccinated; 3% among 3-dose vaccinated). Among IC adults, 2-dose mRNA VE was 72% (95% CI: 69-75) and 3-dose mRNA VE was 81% (95% CI: 76-86) against SARS-CoV-2 infection at hospitalization, compared with 87% (95% CI: 86-87) and 96% (95% CI: 95-97), respectively, among non-IC adults. VE against hospitalization for 2 and 3 doses was significantly lower among IC versus non-IC patients (tests of interaction, both p<0.05). VE point estimates were similarly lower among IC versus non-IC patients in analyses stratified by age group and race/ethnicity (**Supplementary Tables 4-5**).

**Table 4.**
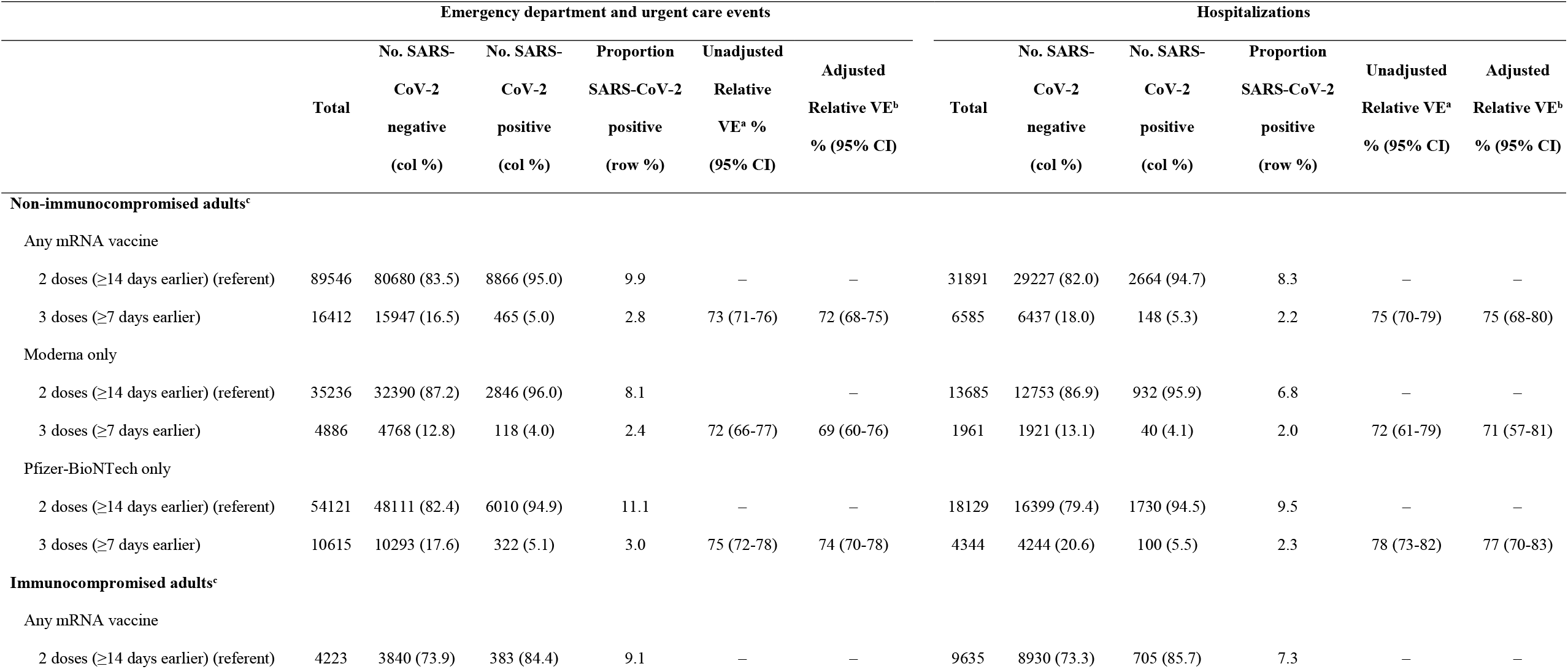
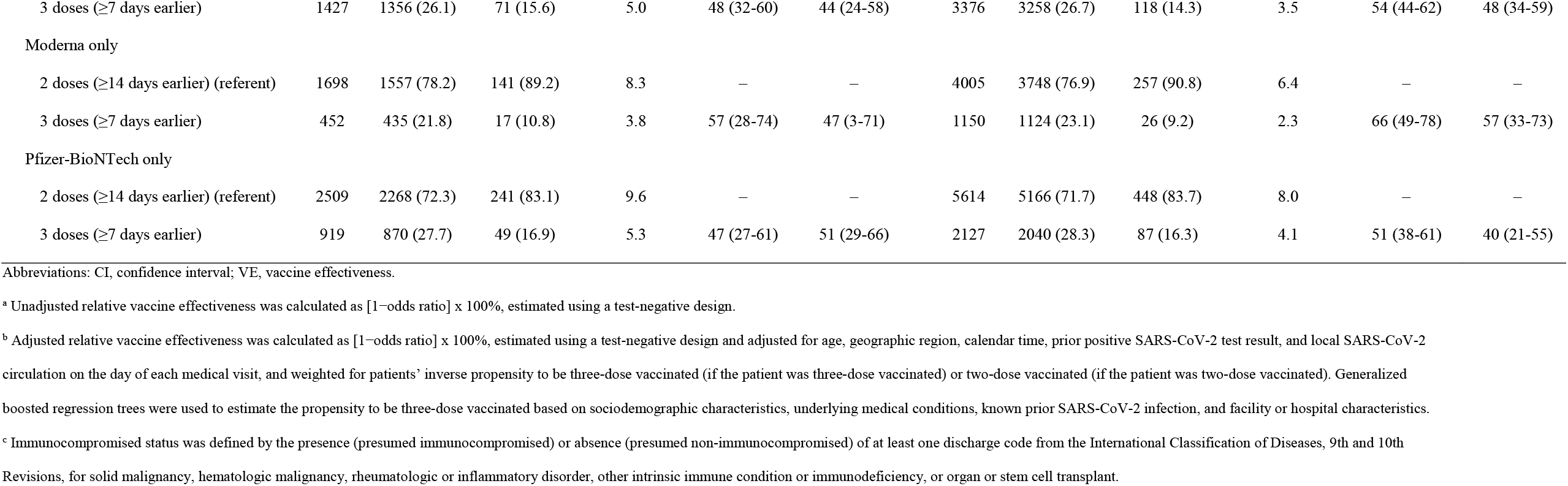
Relative effectiveness of three versus two doses of mRNA COVID-19 vaccines against laboratory-confirmed COVID-19–associated emergency department or urgent care events and hospitalizations among non-immunocompromised and immunocompromised adults, by vaccine product received, August–December 2021.

| Emergency department and urgent care events |  |  |  |  |  |  | Hospitalizations |  |  |  |  |  |
| --- | --- | --- | --- | --- | --- | --- | --- | --- | --- | --- | --- | --- |
|  | Total | No. SARS-CoV-2 negative | No. SARS-CoV-2 positive | Proportion SARS-CoV-2 positive | Unadjusted Relative VE<sup>a</sup> % | Adjusted Relative VE<sup>b</sup> % (95% CI) | Total | No. SARS-CoV-2 negative | No. SARS-CoV-2 positive | Proportion SARS-CoV-2 positive | Unadjusted Relative VE<sup>a</sup> % (95% CI) | Adjusted Relative VE<sup>b</sup> % (95% CI) |
|  |  | (col %) | (col %) | (row %) | (95% CI) |  |  | (col %) | (col %) | (row %) |  |  |
| Non-immunocompromised adults<sup>c</sup> |  |  |  |  |  |  |  |  |  |  |  |  |
| Any mRNA vaccine |  |  |  |  |  |  |  |  |  |  |  |  |
| 2 doses (≥14 days earlier) (referent) | 89546 | 80680 (83.5) | 8866 (95.0) | 9.9 | — | — | 31891 | 29227 (82.0) | 2664 (94.7) | 8.3 | — | — |
| 3 doses (≥7 days earlier) | 16412 | 15947 (16.5) | 465 (5.0) | 2.8 | 73 (71-76) | 72 (68-75) | 6585 | 6437 (18.0) | 148 (5.3) | 2.2 | 75 (70-79) | 75 (68-80) |
| Moderna only |  |  |  |  |  |  |  |  |  |  |  |  |
| 2 doses (≥14 days earlier) (referent) | 35236 | 32390 (87.2) | 2846 (96.0) | 8.1 |  | — | 13685 | 12753 (86.9) | 932 (95.9) | 6.8 | — | — |
| 3 doses (≥7 days earlier) | 4886 | 4768 (12.8) | 118 (4.0) | 2.4 | 72 (66-77) | 69 (60-76) | 1961 | 1921 (13.1) | 40 (4.1) | 2.0 | 72 (61-79) | 71 (57-81) |
| Pfizer-BioNTech only |  |  |  |  |  |  |  |  |  |  |  |  |
| 2 doses (≥14 days earlier) (referent) | 54121 | 48111 (82.4) | 6010 (94.9) | 11.1 | — | — | 18129 | 16399 (79.4) | 1730 (94.5) | 9.5 | — | — |
| 3 doses (≥7 days earlier) | 10615 | 10293 (17.6) | 322 (5.1) | 3.0 | 75 (72-78) | 74 (70-78) | 4344 | 4244 (20.6) | 100 (5.5) | 2.3 | 78 (73-82) | 77 (70-83) |
| Immunocompromised adults<sup>c</sup> |  |  |  |  |  |  |  |  |  |  |  |  |
| Any mRNA vaccine |  |  |  |  |  |  |  |  |  |  |  |  |
| 2 doses (≥14 days earlier) (referent) | 4223 | 3840 (73.9) | 383 (84.4) | 9.1 | — | — | 9635 | 8930 (73.3) | 705 (85.7) | 7.3 | — | — |
| 3 doses (≥7 days earlier) | 1427 | 1356 (26.1) | 71 (15.6) | 5.0 | 48 (32-60) | 44 (24-58) | 3376 | 3258 (26.7) | 118 (14.3) | 3.5 | 54 (44-62) | 48 (34-59) |
| Moderna only |  |  |  |  |  |  |  |  |  |  |  |  |
| 2 doses (≥14 days earlier) (referent) | 1698 | 1557 (78.2) | 141 (89.2) | 8.3 | — | — | 4005 | 3748 (76.9) | 257 (90.8) | 6.4 | — | — |
| 3 doses (≥7 days earlier) | 452 | 435 (21.8) | 17 (10.8) | 3.8 | 57 (28-74) | 47 (3-71) | 1150 | 1124 (23.1) | 26 (9.2) | 2.3 | 66 (49-78) | 57 (33-73) |
| Pfizer-BioNTech only |  |  |  |  |  |  |  |  |  |  |  |  |
| 2 doses (≥14 days earlier) (referent) | 2509 | 2268 (72.3) | 241 (83.1) | 9.6 | — | — | 5614 | 5166 (71.7) | 448 (83.7) | 8.0 | — | — |
| 3 doses (≥7 days earlier) | 919 | 870 (27.7) | 49 (16.9) | 5.3 | 47 (27-61) | 51 (29-66) | 2127 | 2040 (28.3) | 87 (16.3) | 4.1 | 51 (38-61) | 40 (21-55) |

Abbreviations: CI, confidence interval; VE, vaccine effectiveness.
<sup>a</sup> Unadjusted relative vaccine effectiveness was calculated as [1–odds ratio] x 100%, estimated using a test-negative design.
<sup>b</sup> Adjusted relative vaccine effectiveness was calculated as [1–odds ratio] x 100%, estimated using a test-negative design and adjusted for age, geographic region, calendar time, prior positive SARS-CoV-2 test result, and local SARS-CoV-2 circulation on the day of each medical visit, and weighted for patients’ inverse propensity to be three-dose vaccinated (if the patient was three-dose vaccinated) or two-dose vaccinated (if the patient was two-dose vaccinated). Generalized boosted regression trees were used to estimate the propensity to be three-dose vaccinated based on sociodemographic characteristics, underlying medical conditions, known prior SARS-CoV-2 infection, and facility or hospital characteristics.
<sup>c</sup> Immunocompromised status was defined by the presence (presumed immunocompromised) or absence (presumed non-immunocompromised) of at least one discharge code from the International Classification of Diseases, 9th and 10th Revisions, for solid malignancy, hematologic malignancy, rheumatologic or inflammatory disorder, other intrinsic immune condition or immunodeficiency, or organ or stem cell transplant.

In ED/UC and hospital settings, point estimates for 2-dose mRNA VE were lower among patients with ≥150 versus <150 days elapsed since second dose (**Table 3**). Further, among IC patients in both settings, 2-dose VE was higher for Moderna than Pfizer-BioNTech products (both p<0.05). Similarly, among IC patients, 3-dose VE was higher for Moderna than Pfizer-BioNTech products in hospital settings (p<0.05), but no significant difference was observed in ED/UC settings (p=0.61). In relative VE analyses among IC patients, three mRNA doses were significantly more effective than two mRNA doses in ED/UC settings (44%; 95% CI: 24–58) and hospital settings (48%; 95% CI: 34–59); results were similar for both Moderna and Pfizer-BioNTech products (**Table 4)**.

### mRNA COVID-19 VE by Type of Immunocompromising Condition

Among patients with only one type of immunocompromising condition, VE against COVID-19– associated ED/UC events after 2-dose mRNA vaccination for IC patient groups ranged from 11% (95% CI: −102-61) among organ/stem cell transplant recipients to 82% (95% CI: 66-90) among patients with hematologic malignancy. 3-dose VE ranged from 50% (95% CI: −36-81) among patients with hematologic malignancy to 87% (95% CI: 70-95) among patients with solid malignancy (**Table 5**). VE against COVID-19–associated hospitalizations of patients with 2-dose mRNA vaccination ranged from 73% (95% CI: 49-86) among organ/stem cell transplant recipients to 83% (95% CI: 77-87) among patients with another intrinsic immune condition or underlying immunodeficiency. VE of 3-dose mRNA vaccination ranged from 39% (95% CI: −69-78) among organ/stem cell transplant recipients to 92% (95% CI: 84-96) among patients with solid malignancy.

**Table 5.**
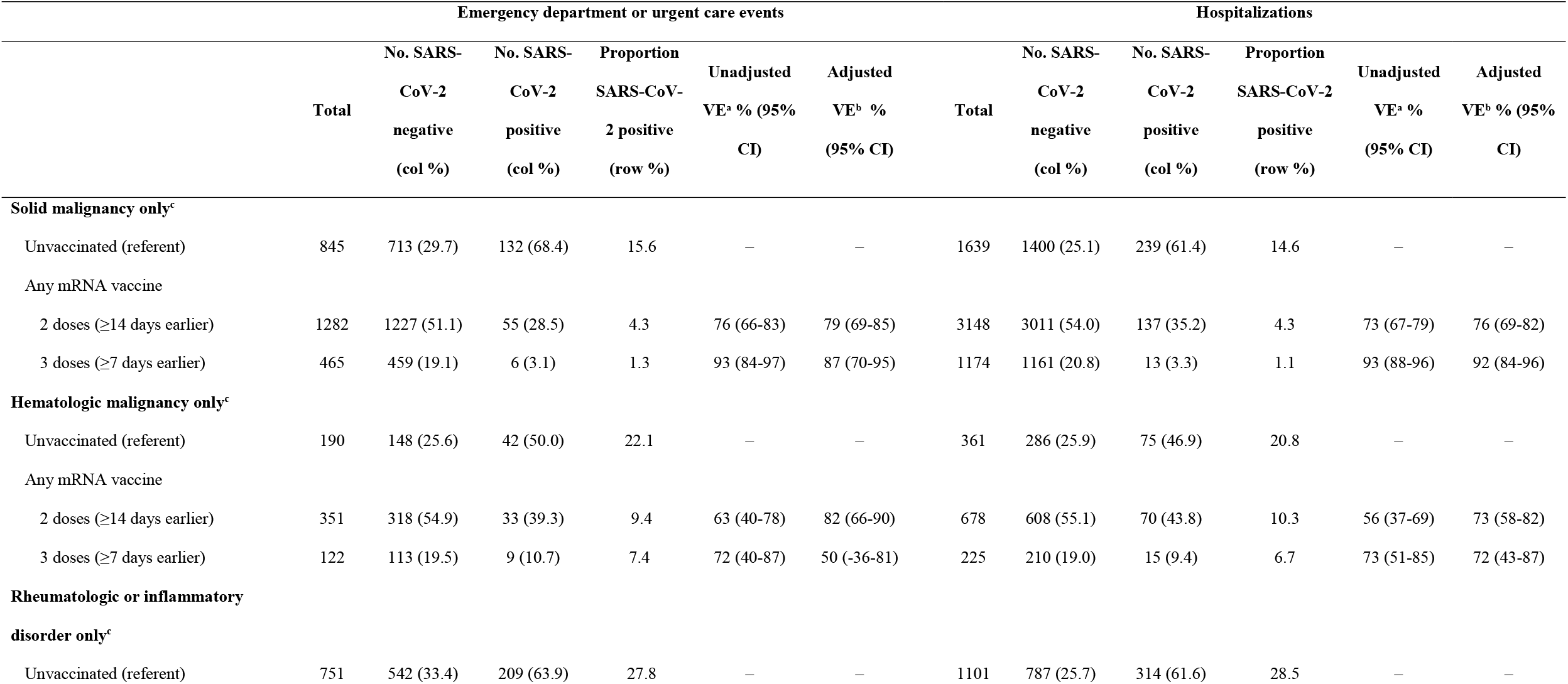
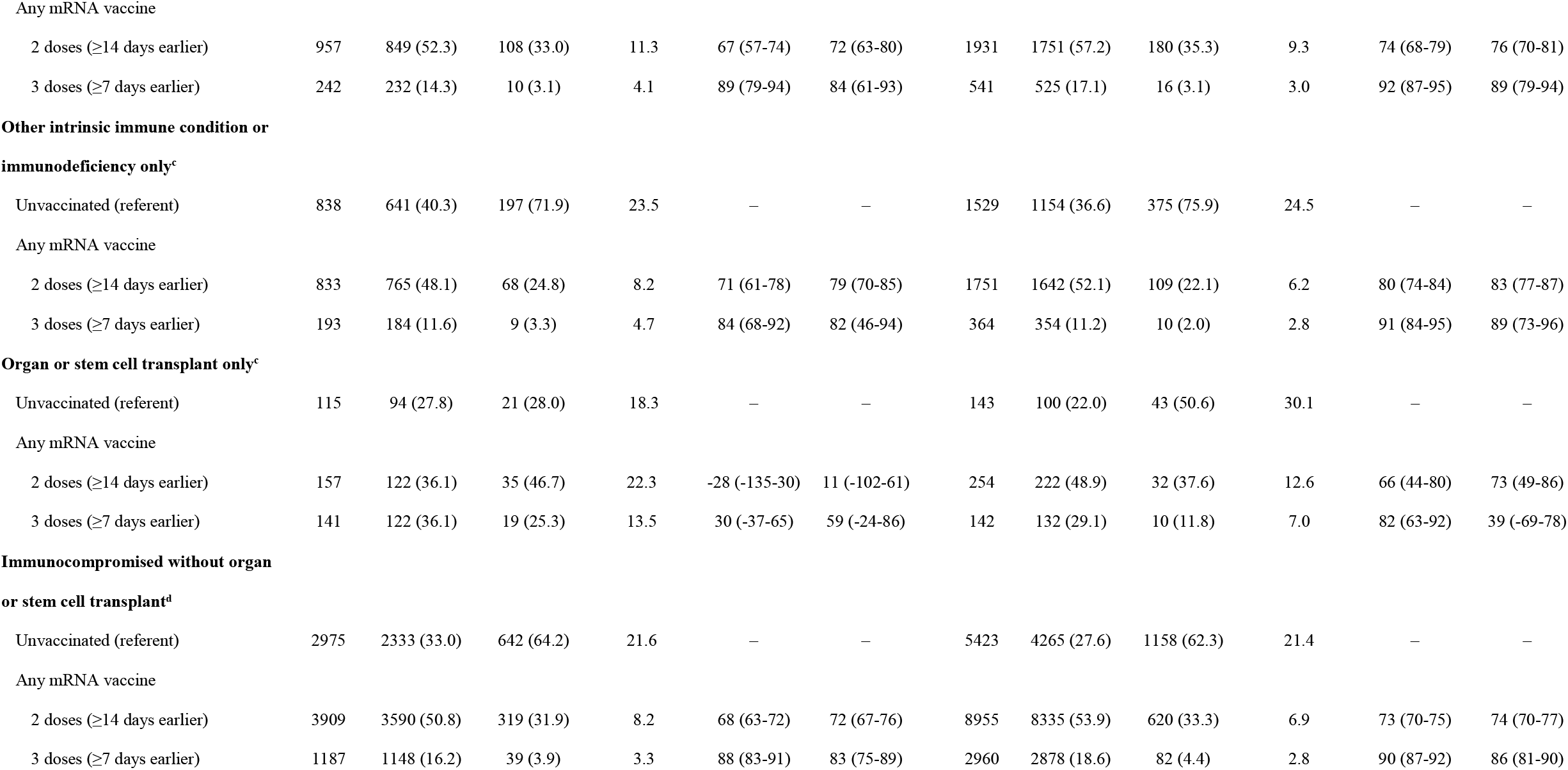
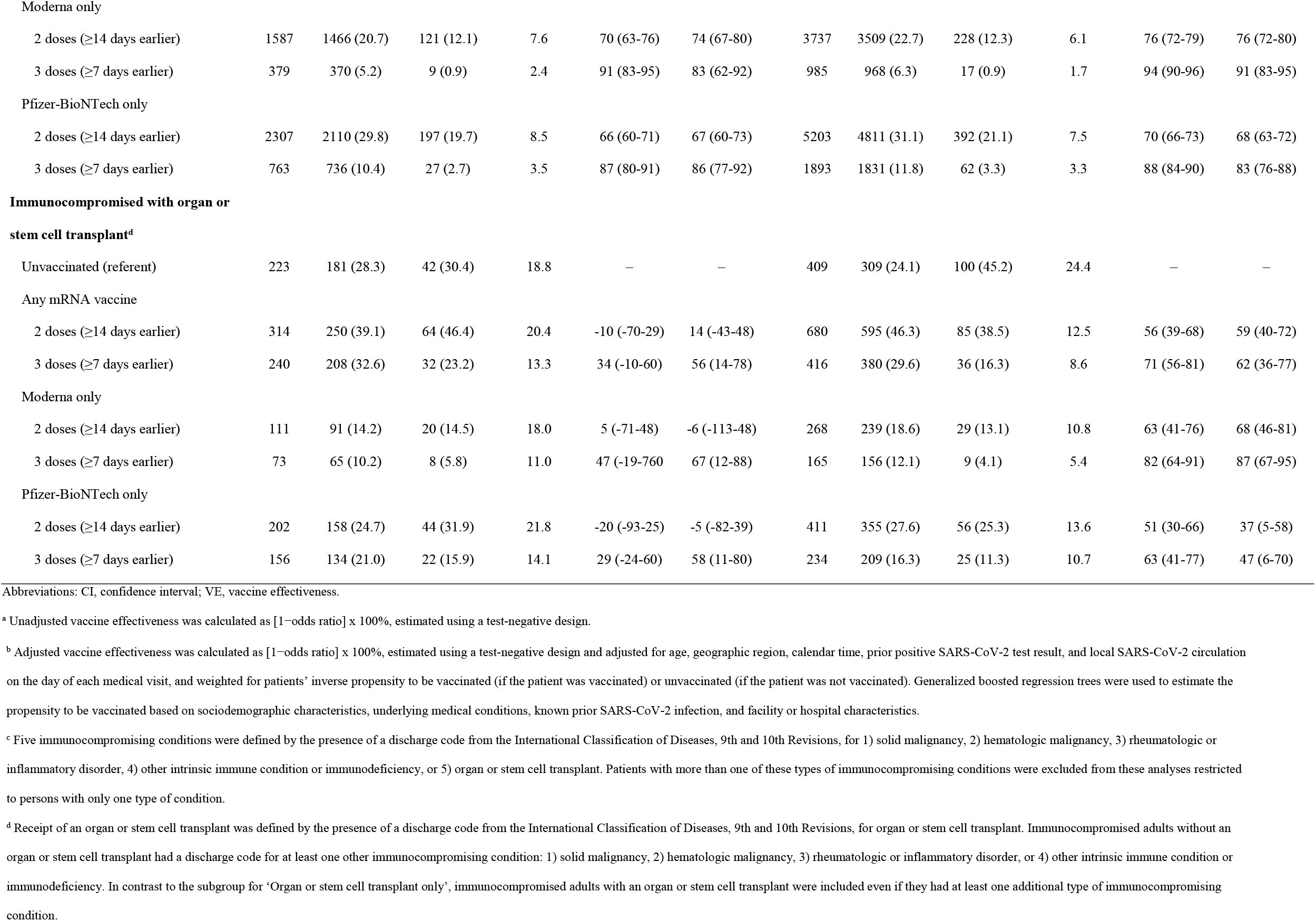
mRNA COVID-19 vaccine effectiveness against laboratory-confirmed COVID-19–associated emergency department or urgent care events and hospitalizations among subgroups of adults with specific types of immunocompromising conditions, by number of vaccine doses, August–December 2021.

| Emergency department or urgent care events |  |  |  |  |  |  | Hospitalizations |  |  |  |  |  |
| --- | --- | --- | --- | --- | --- | --- | --- | --- | --- | --- | --- | --- |
|  | Total | No. SARS-CoV-2 negative | No. SARS-CoV-2 positive | Proportion SARS-CoV-2 positive | Unadjusted VE<sup>a</sup> % (95% CI) | Adjusted VE<sup>b</sup> % (95% CI) | Total | No. SARS-CoV-2 negative | No. SARS-CoV-2 positive | Proportion SARS-CoV-2 positive | Unadjusted VE<sup>a</sup> % (95% CI) | Adjusted VE<sup>b</sup> % (95% CI) |
|  |  | (col %) | (col %) | (row %) |  |  |  | (col %) | (col %) | (row %) |  |  |
| Solid malignancy only<sup>c</sup> |  |  |  |  |  |  |  |  |  |  |  |  |
| Unvaccinated (referent) | 845 | 713 (29.7) | 132 (68.4) | 15.6 | — | — | 1639 | 1400 (25.1) | 239 (61.4) | 14.6 | — | — |
| Any mRNA vaccine |  |  |  |  |  |  |  |  |  |  |  |  |
| 2 doses (≥14 days earlier) | 1282 | 1227 (51.1) | 55 (28.5) | 4.3 | 76 (66-83) | 79 (69-85) | 3148 | 3011 (54.0) | 137 (35.2) | 4.3 | 73 (67-79) | 76 (69-82) |
| 3 doses (≥7 days earlier) | 465 | 459 (19.1) | 6 (3.1) | 1.3 | 93 (84-97) | 87 (70-95) | 1174 | 1161 (20.8) | 13 (3.3) | 1.1 | 93 (88-96) | 92 (84-96) |
| Hematologic malignancy only<sup>c</sup> |  |  |  |  |  |  |  |  |  |  |  |  |
| Unvaccinated (referent) | 190 | 148 (25.6) | 42 (50.0) | 22.1 | — | — | 361 | 286 (25.9) | 75 (46.9) | 20.8 | — | — |
| Any mRNA vaccine |  |  |  |  |  |  |  |  |  |  |  |  |
| 2 doses (≥14 days earlier) | 351 | 318 (54.9) | 33 (39.3) | 9.4 | 63 (40-78) | 82 (66-90) | 678 | 608 (55.1) | 70 (43.8) | 10.3 | 56 (37-69) | 73 (58-82) |
| 3 doses (≥7 days earlier) | 122 | 113 (19.5) | 9 (10.7) | 7.4 | 72 (40-87) | 50 (-36-81) | 225 | 210 (19.0) | 15 (9.4) | 6.7 | 73 (51-85) | 72 (43-87) |
| Rheumatologic or inflammatory disorder only<sup>c</sup> |  |  |  |  |  |  |  |  |  |  |  |  |
| Unvaccinated (referent) | 751 | 542 (33.4) | 209 (63.9) | 27.8 | — | — | 1101 | 787 (25.7) | 314 (61.6) | 28.5 | — | — |
| Any mRNA vaccine |  |  |  |  |  |  |  |  |  |  |  |  |
| 2 doses (≥14 days earlier) | 957 | 849 (52.3) | 108 (33.0) | 11.3 | 67 (57-74) | 72 (63-80) | 1931 | 1751 (57.2) | 180 (35.3) | 9.3 | 74 (68-79) | 76 (70-81) |
| 3 doses (≥7 days earlier) | 242 | 232 (14.3) | 10 (3.1) | 4.1 | 89 (79-94) | 84 (61-93) | 541 | 525 (17.1) | 16 (3.1) | 3.0 | 92 (87-95) | 89 (79-94) |
| **Other intrinsic immune condition or immunodeficiency only<sup>c</sup>** |  |  |  |  |  |  |  |  |  |  |  |  |
| Unvaccinated (referent) | 838 | 641 (40.3) | 197 (71.9) | 23.5 | — | — | 1529 | 1154 (36.6) | 375 (75.9) | 24.5 | — | — |
| Any mRNA vaccine |  |  |  |  |  |  |  |  |  |  |  |  |
| 2 doses (≥14 days earlier) | 833 | 765 (48.1) | 68 (24.8) | 8.2 | 71 (61-78) | 79 (70-85) | 1751 | 1642 (52.1) | 109 (22.1) | 6.2 | 80 (74-84) | 83 (77-87) |
| 3 doses (≥7 days earlier) | 193 | 184 (11.6) | 9 (3.3) | 4.7 | 84 (68-92) | 82 (46-94) | 364 | 354 (11.2) | 10 (2.0) | 2.8 | 91 (84-95) | 89 (73-96) |
| **Organ or stem cell transplant only<sup>c</sup>** |  |  |  |  |  |  |  |  |  |  |  |  |
| Unvaccinated (referent) | 115 | 94 (27.8) | 21 (28.0) | 18.3 | — | — | 143 | 100 (22.0) | 43 (50.6) | 30.1 | — | — |
| Any mRNA vaccine |  |  |  |  |  |  |  |  |  |  |  |  |
| 2 doses (≥14 days earlier) | 157 | 122 (36.1) | 35 (46.7) | 22.3 | -28 (-135-30) | 11 (-102-61) | 254 | 222 (48.9) | 32 (37.6) | 12.6 | 66 (44-80) | 73 (49-86) |
| 3 doses (≥7 days earlier) | 141 | 122 (36.1) | 19 (25.3) | 13.5 | 30 (-37-65) | 59 (-24-86) | 142 | 132 (29.1) | 10 (11.8) | 7.0 | 82 (63-92) | 39 (-69-78) |
| **Immunocompromised without organ or stem cell transplant<sup>d</sup>** |  |  |  |  |  |  |  |  |  |  |  |  |
| Unvaccinated (referent) | 2975 | 2333 (33.0) | 642 (64.2) | 21.6 | — | — | 5423 | 4265 (27.6) | 1158 (62.3) | 21.4 | — | — |
| Any mRNA vaccine |  |  |  |  |  |  |  |  |  |  |  |  |
| 2 doses (≥14 days earlier) | 3909 | 3590 (50.8) | 319 (31.9) | 8.2 | 68 (63-72) | 72 (67-76) | 8955 | 8335 (53.9) | 620 (33.3) | 6.9 | 73 (70-75) | 74 (70-77) |
| 3 doses (≥7 days earlier) | 1187 | 1148 (16.2) | 39 (3.9) | 3.3 | 88 (83-91) | 83 (75-89) | 2960 | 2878 (18.6) | 82 (4.4) | 2.8 | 90 (87-92) | 86 (81-90) |
| Moderna only |  |  |  |  |  |  |  |  |  |  |  |  |
| 2 doses (≥14 days earlier) | 1587 | 1466 (20.7) | 121 (12.1) | 7.6 | 70 (63-76) | 74 (67-80) | 3737 | 3509 (22.7) | 228 (12.3) | 6.1 | 76 (72-79) | 76 (72-80) |
| 3 doses (≥7 days earlier) | 379 | 370 (5.2) | 9 (0.9) | 2.4 | 91 (83-95) | 83 (62-92) | 985 | 968 (6.3) | 17 (0.9) | 1.7 | 94 (90-96) | 91 (83-95) |
| Pfizer-BioNTech only |  |  |  |  |  |  |  |  |  |  |  |  |
| 2 doses (≥14 days earlier) | 2307 | 2110 (29.8) | 197 (19.7) | 8.5 | 66 (60-71) | 67 (60-73) | 5203 | 4811 (31.1) | 392 (21.1) | 7.5 | 70 (66-73) | 68 (63-72) |
| 3 doses (≥7 days earlier) | 763 | 736 (10.4) | 27 (2.7) | 3.5 | 87 (80-91) | 86 (77-92) | 1893 | 1831 (11.8) | 62 (3.3) | 3.3 | 88 (84-90) | 83 (76-88) |
| **Immunocompromised with organ or stem cell transplant<sup>a</sup>** |  |  |  |  |  |  |  |  |  |  |  |  |
| Unvaccinated (referent) | 223 | 181 (28.3) | 42 (30.4) | 18.8 | — | — | 409 | 309 (24.1) | 100 (45.2) | 24.4 | — | — |
| Any mRNA vaccine |  |  |  |  |  |  |  |  |  |  |  |  |
| 2 doses (≥14 days earlier) | 314 | 250 (39.1) | 64 (46.4) | 20.4 | -10 (-70-29) | 14 (-43-48) | 680 | 595 (46.3) | 85 (38.5) | 12.5 | 56 (39-68) | 59 (40-72) |
| 3 doses (≥7 days earlier) | 240 | 208 (32.6) | 32 (23.2) | 13.3 | 34 (-10-60) | 56 (14-78) | 416 | 380 (29.6) | 36 (16.3) | 8.6 | 71 (56-81) | 62 (36-77) |
| Moderna only |  |  |  |  |  |  |  |  |  |  |  |  |
| 2 doses (≥14 days earlier) | 111 | 91 (14.2) | 20 (14.5) | 18.0 | 5 (-71-48) | -6 (-113-48) | 268 | 239 (18.6) | 29 (13.1) | 10.8 | 63 (41-76) | 68 (46-81) |
| 3 doses (≥7 days earlier) | 73 | 65 (10.2) | 8 (5.8) | 11.0 | 47 (-19-760) | 67 (12-88) | 165 | 156 (12.1) | 9 (4.1) | 5.4 | 82 (64-91) | 87 (67-95) |
| Pfizer-BioNTech only |  |  |  |  |  |  |  |  |  |  |  |  |
| 2 doses (≥14 days earlier) | 202 | 158 (24.7) | 44 (31.9) | 21.8 | -20 (-93-25) | -5 (-82-39) | 411 | 355 (27.6) | 56 (25.3) | 13.6 | 51 (30-66) | 37 (5-58) |
| 3 doses (≥7 days earlier) | 156 | 134 (21.0) | 22 (15.9) | 14.1 | 29 (-24-60) | 58 (11-80) | 234 | 209 (16.3) | 25 (11.3) | 10.7 | 63 (41-77) | 47 (6-70) |

Abbreviations: CI, confidence interval; VE, vaccine effectiveness.
<sup>a</sup> Unadjusted vaccine effectiveness was calculated as [1–odds ratio] x 100%, estimated using a test-negative design.
<sup>b</sup> Adjusted vaccine effectiveness was calculated as $[1 - \text{odds ratio}] \times 100\%$ , estimated using a test-negative design and adjusted for age, geographic region, calendar time, prior positive SARS-CoV-2 test result, and local SARS-CoV-2 circulation on the day of each medical visit, and weighted for patients' inverse propensity to be vaccinated (if the patient was vaccinated) or unvaccinated (if the patient was not vaccinated). Generalized boosted regression trees were used to estimate the propensity to be vaccinated based on sociodemographic characteristics, underlying medical conditions, known prior SARS-CoV-2 infection, and facility or hospital characteristics.
<sup>c</sup> Five immunocompromising conditions were defined by the presence of a discharge code from the International Classification of Diseases, 9th and 10th Revisions, for 1) solid malignancy, 2) hematologic malignancy, 3) rheumatologic or inflammatory disorder, 4) other intrinsic immune condition or immunodeficiency, or 5) organ or stem cell transplant. Patients with more than one of these types of immunocompromising conditions were excluded from these analyses restricted to persons with only one type of condition.
<sup>d</sup> Receipt of an organ or stem cell transplant was defined by the presence of a discharge code from the International Classification of Diseases, 9th and 10th Revisions, for organ or stem cell transplant. Immunocompromised adults without an organ or stem cell transplant had a discharge code for at least one other immunocompromising condition: 1) solid malignancy, 2) hematologic malignancy, 3) rheumatologic or inflammatory disorder, or 4) other intrinsic immune condition or immunodeficiency. In contrast to the subgroup for 'Organ or stem cell transplant only', immunocompromised adults with an organ or stem cell transplant were included even if they had at least one additional type of immunocompromising condition.

VE of 2-dose and 3-dose mRNA vaccination (each versus unvaccinated status) was lower among transplant recipients than among non-transplant recipients with other immunocompromising condition(s) in ED/UC and inpatient settings (tests of interaction between organ/stem cell transplant status and vaccination status, all p<0.05) (**Table 5**). Among transplant recipients, two or three doses of Moderna were more effective than two or three doses of Pfizer-BioNTech against COVID-19 hospitalization (both p<0.05), but no product difference was detected with respect to VE against COVID-19 ED/UC events (both p>0.25).

### Janssen (Johnson & Johnson) COVID-19 VE

A total of 10,329 ED/UC events (5% IC) and 4,202 hospitalizations (23% IC) were among Janssen vaccine recipients. Characteristics of non-IC and IC Janssen vaccine recipients in ED/UC and hospital settings are provided in **Supplementary Tables 6-7**. Among Janssen vaccine recipients, 7% of non-IC patients and 6% of IC patients in ED/UC settings, and 6% and 5%, respectively, in hospital settings, had received a second dose (any product). Among those who received a second dose, 67% and 63% of non-IC patients in ED/UC and hospital settings, respectively, and 77% and 69% of IC patients in those settings had a heterologous dosing pattern (i.e., mRNA product as second dose).

In ED/UC settings, median duration since 1-dose Janssen-vaccinated patients’ first dose was 182 days among non-IC patients (IQR: 139–219) and 190 days among IC patients (IQR: 148–227); median durations in hospitals were 179 days (IQR: 134–215) and 185 days (IQR: 147–218), respectively. In ED/UC settings, median duration since 2-dose Janssen-vaccinated patients’ second dose (any product) was 26 days among non-IC patients (IQR: 16-40) and 20 days among IC patients (IQR: 12–37); median durations in hospitals were 26 days (IQR: 15–38) and 29 days (IQR: 17–40), respectively.

Among IC adults, VE of one Janssen dose was 7% (95% CI: −26–31) and VE of two doses (any product for second dose) was 71% (95% CI: 6–91) against COVID-19 ED/UC events, compared with 64% (95% CI: 61–66) and 90% (95% CI: 86–93), respectively, among non-IC adults (**see Supplementary Table 8**). VE for 1 Janssen dose was significantly lower among IC versus non-IC patients (test of interaction, p<0.05); there was no statistical difference for 2 doses. Among IC adults, VE of one Janssen dose was 45% (CI: 29–57) and VE of two doses was 70% (95% CI: 25–88) against COVID-19 hospitalization, compared with 72% (95% CI: 69–76) and 93% (95% CI: 86–96), respectively, among non-IC adults (tests of interaction between IC status and vaccination status, p<0.05 for 1 dose and 2 doses).

## DISCUSSION

In a multistate analysis of real-world EHR-derived data from nearly 209,000 ED/UC events and 90,000 hospitalizations during Delta variant predominance, 3 doses among mRNA COVID-19 vaccine recipients and 2 doses among viral-vector vaccine recipients (any product for second dose) were more effective at preventing laboratory-confirmed COVID-19 medical events in IC and non-IC patients compared with receipt of 2 mRNA vaccine doses, one viral-vector vaccine dose, or being unvaccinated. While this additional primary series dose offers further protection to IC patients, VE estimates among IC patients who received 3 doses of mRNA vaccines or 2 doses of a viral-vector containing series were significantly lower than VE among non-IC patients. These findings indicate that, as a group, IC patients receive less protection from vaccination, compared with the protection received by non-IC patients. These findings are consistent with other studies suggesting that certain IC populations experience attenuated immune responses to COVID-19 vaccines and that VE is reduced among IC patients overall [3, 27]. To our knowledge, this study is among the first to report different estimates for 3-dose mRNA VE across groups with different IC conditions, with point estimates ranging from 50–87% (ED/UC) and 39– 92% (hospital). Specifically, and consistent with our previous study of 2-dose vaccination, VE was again found to be especially low for patients with organ or stem cell transplants, confirming that this population is at particular risk of lower protection from severe COVID-19 outcomes even when vaccinated [7].

Our results highlight the need for extra protection against COVID-19 among IC adults. Therefore, they support the recommendation that all eligible persons should receive multiple vaccine doses, and that IC persons should receive more than non-IC persons, to enhance protection against severe COVID-19. They also add to evidence suggesting that, compared with non-IC persons, IC populations may have a greater need for additional measures (masking, physical distancing, pre-exposure prophylaxis) to prevent COVID-19.

To our knowledge, this study is also among the first to show a statistically significant difference in VE between mRNA products among IC patients. Our network’s previous study demonstrated a trend toward a greater protective effect of the Moderna versus Pfizer-BioNTech mRNA vaccine, and this study provides more rigorous statistical evidence of this difference [7]. Therefore, while this is an observational study with some inherent limitations noted below, and while either mRNA vaccine provides good VE for IC patients, this finding is notable and deserving of further study and consideration. It should also be noted that we did not have sufficient power to directly compare relative VE between viral-vector and mRNA vaccine products.

This study has several strengths, including its large and diverse sample, but it is also subject to several limitations. First, the use of selected discharge diagnoses as surrogates for presumed IC status and the absence of medication and other relevant data might have led to classification of persons as IC who were not; the opposite is also possible but less likely. Second, despite each network partner’s data linkage with state or local immunization registries, misclassification of vaccination status is possible. Third, although inverse weights balanced unvaccinated and vaccinated patients on sociodemographic and health characteristics, and further adjustments were made, unmeasured and residual confounding (e.g., mask-wearing, waning immunity) in this observational study might have influenced estimates. For example, prior SARS-CoV-2 infection could only be controlled for if it was documented in the EHR. Fourth, this study was limited to the period of Delta variant predominance and findings may differ during the subsequent period of Omicron variant predominance, a matter that will be the focus of future analyses. Finally, although additional mRNA doses are currently recommended for IC patients, current data availability did not yet allow for such analyses, and will also be the focus of future studies.

This study has implications for public health and clinical practice. While further analyses should evaluate the effectiveness of four mRNA vaccine doses, patients will benefit from receiving booster COVID-19 vaccine doses. Consistent with current CDC recommendations, booster doses confer greater protection than the primary series alone [8]. However, this study also reinforces the reality that vaccinated IC patients remain less protected than vaccinated non-IC patients and may benefit more from additional precautions, including the use of nonpharmaceutical prevention strategies such as mask-wearing to prevent SARS-CoV-2 infection. Also, if infected with SARS-CoV-2, IC patients would benefit from being monitored closely and considered early for proven therapies that might prevent progression to severe illness. Additional studies are needed to determine the degree of VE improvement conferred by additional COVID-19 vaccine doses in IC populations, evaluate whether different approaches to vaccine administration might improve VE (e.g., dosage timing, temporarily withholding immunosuppressants), and further evaluate VE differences between vaccines products.

## Data Availability

The individual level dataset from this study is held securely in limited deidentified form at the US Centers for Disease Control and Prevention. Data sharing agreements between CDC and data providers prohibit CDC from making this dataset publicly available. CDC will share aggregate study data once study objectives are complete, consistent with data use agreements with partner institutions.

## Acknowledgements

We would like to acknowledge the contributions of the VISION Network beyond the authors listed; Baylor Scott & White Health: Deborah Hendricks, Jason Ettlinger, Joel Blais, Elisa Priest, Michael Smith, Spencer Rose, Natalie Settele, Jennifer Thomas, Muralidhar Jatla, Madhava Beeram, Alejandro Arroliga; CDC: Catherine H. Bozio, Jill Ferdinands, Eric P. Griggs, Rachael M. Porter, Lenee Blanton, Andrea Steffens, Natalie Olson, Jeremiah Williams, Monica Dickerson, Meredith McMorrow, Stephanie J. Schrag, Jennifer R. Verani, Mehiret Wondimu; Center for Health Research, Kaiser Permanente Northwest: Padma Dandamudi, Bradley Crane; HealthPartners: Gabriela Vazquez-Benitez, Inih Essien, Sunita Thapa, Sheryl Kane; School of Medicine, University of Colorado Anschutz Medical Campus: Health Data Compass, David Mayer, Bryant Doyle, Briana Kille, Catia Chavez; Regenstrief Institute: Ashley Wiensch, Amy Hancock; Westat: Maria Demarco, Salome Kiduko, Carly Hallowell, Magdalene Kish, Lemlem Mehari, Yan Zhuang, Alex Hughes, Kristen Butterfield, Jack Davis, Patrick Mitchell, Victoria Lazariu, Weijia Ren, Akintunde Akinseye.

## Patient Consent Statement

This study was reviewed and approved by the institutional review board at Westat, Inc. This electronic health record-based study does not include factors necessitating patient consent.

## References

1. Williamson EJ, Walker AJ, Bhaskaran K, et al. Factors associated with COVID-19-related death using OpenSAFELY. Nature 2020; 584(7821):430–6.

2. Harpaz R, Dahl RM, Dooling KL. Prevalence of immunosuppression among US adults, 2013. JAMA 2016; 316(23):2547.

3. Deepak P, Kim W, Paley MA, et al. Effect of immunosuppression on the immunogenicity of mRNA vaccines to SARS-CoV-2: a prospective cohort study. Ann Intern Med 2021; 174(11):1572–85.

4. Negahdaripour M, Shafiekhani M, Moezzi SMI, et al. Administration of COVID-19 vaccines in immunocompromised patients. Int Immunopharmacol 2021; 99:108021.

5. Di Fusco M, Lin J, Vaghela S, et al. COVID-19 vaccine effectiveness among immunocompromised populations: a targeted literature review of real-world studies. Expert Rev Vaccines. 2022; 21(4):435–51.

6. Haidar G, Agha M, Bilderback A, et al. Prospective evaluation of COVID-19 vaccine responses across a broad spectrum of immunocompromising conditions: the COVICS study. Clin Infect Dis 2022; 75(1):e630–e644.

7. Embi PJ, Levy ME, Naleway AL, et al. Effectiveness of 2-dose vaccination with mRNA COVID-19 vaccines against COVID-19–associated hospitalizations among immunocompromised adults — nine states, January–September 2021. MMWR Morb Mortal Wkly Rep 2021; 70(44):1553–9.

8. Centers for Disease Control and Prevention. COVID-19 vaccines for moderately or severely immunocompromised people 2022 [Available from: https://www.cdc.gov/coronavirus/2019-ncov/vaccines/recommendations/immuno.html#:~:text=decreased%20over%20time.-,People%20ages%2012%20years%20and%20older%20who%20are%20moderately%20or,when%20eligible%2C%201%20booster%20dose].

9. Tenforde MW, Patel MM, Gaglani M, et al. Effectiveness of a third dose of Pfizer-BioNTech and Moderna vaccines in preventing COVID-19 hospitalization among immunocompetent and immunocompromised adults - United States, August-December 2021. MMWR Morb Mortal Wkly Rep 2022; 71(4):118–24.

10. Kwon JH, Tenforde MW, Gaglani M, et al. mRNA vaccine effectiveness against COVID-19 hospitalization among solid organ transplant recipients. J Infect Dis 2022; 226(5):797–807.

11. Thompson MG, Stenehjem E, Grannis S, et al. Effectiveness of Covid-19 vaccines in ambulatory and inpatient care settings. N Engl J Med 2021; 385(15):1355–71.

12. Grannis SJ, Rowley EA, Ong TC, et al. Interim estimates of COVID-19 vaccine effectiveness against COVID-19–associated emergency department or urgent care clinic encounters and hospitalizations among adults during SARS-CoV-2 B.1.617.2 (Delta) variant predominance — nine states, June– August 2021. MMWR Morb Mortal Wkly Rep 2021; 70(37):1291–3.

13. Thompson MG, Natarajan K, Irving SA, et al. Effectiveness of a third dose of mRNA vaccines against COVID-19–associated emergency department and urgent care encounters and hospitalizations among adults during periods of Delta and Omicron variant predominance - VISION Network, 10 states, August 2021-January 2022. MMWR Morb Mortal Wkly Rep 2022; 71(4):139–45.

14. Ferdinands JM, Rao S, Dixon BE, et al. Waning 2-dose and 3-dose effectiveness of mRNA vaccines against COVID-19–associated emergency department and urgent care encounters and hospitalizations among adults during periods of Delta and Omicron variant predominance - VISION Network, 10 states, August 2021-January 2022. MMWR Morb Mortal Wkly Rep 2022; 71(7):255–63.

15. Klein NP, Stockwell MS, Demarco M, et al. Effectiveness of COVID-19 Pfizer-BioNTech BNT162b2 mRNA vaccination in preventing COVID-19-associated emergency department and urgent care encounters and hospitalizations among non-immunocompromised children and adolescents aged 5-17 Years - VISION Network, 10 states, April 2021-January 2022. MMWR Mor Mortal Wkly Rep 2022; 71(9):352–8.

16. Dean NE, Hogan JW, Schnitzer ME. COVID-19 vaccine effectiveness and the test-negative design. N Engl J Med 2021; 385(15):1431–3.

17. Vandenbroucke JP, Pearce N. Test-negative designs: differences and commonalities with other casecontrol studies with “other patient” controls. Epidemiology 2019; 30(6):838–44.

18. Sullivan SG, Tchetgen Tchetgen EJ, Cowling BJ. Theoretical basis of the test-negative study design for assessment of Influenza vaccine effectiveness. Am J Epidemiol 2016; 184(5):345–53.

19. Jackson ML, Nelson JC. The test-negative design for estimating influenza vaccine effectiveness. Vaccine 2013; 31(17):2165–8.

20. Garg S, Kim L, Whitaker M, et al. Hospitalization rates and characteristics of patients hospitalized with laboratory-confirmed Coronavirus disease 2019 - COVID-NET, 14 states, March 1-30, 2020. MMWR Morb Mortal Wkly Rep 2020; 69(15):458–64.

21. Kim L, Garg S, O’Halloran A, et al. Risk Factors for intensive care unit admission and in-hospital mortality among hospitalized adults identified through the US Coronavirus Disease 2019 (COVID-19)-associated hospitalization surveillance network (COVID-NET). Clin Infect Dis 2021; 72(9):e206–e14.

22. Cates J, Lucero-Obusan C, Dahl RM, et al. Risk for in-hospital complications associated with COVID-19 and Influenza - veterans health administration, United States, October 1, 2018-May 31, 2020. MMWR Morb Mortal Wkly Rep 2020; 69(42):1528–34.

23. U.S. Food and Drug Administration. Coronavirus (COVID-19) update: FDA authorizes additional vaccine dose for certain immunocompromised individuals 2021 [Available from: https://www.fda.gov/news-events/press-announcements/coronavirus-covid-19-update-fda-authorizes-additional-vaccine-dose-certain-immunocompromised].

24. Greenberg JA, Hohmann SF, Hall JB, et al. Validation of a method to identify immunocompromised patients with severe sepsis in administrative databases. Ann Am Thorac Soc 2016; 13(2):253–8.

25. Hughes K, Middleton DB, Nowalk MP, et al. Effectiveness of influenza vaccine for preventing laboratory-confirmed Influenza hospitalizations in immunocompromised adults. Clin Infect Dis 2021; 73(11):e4353–e60.

26. Patel M, Chen J, Kim S, et al. Analysis of MarketScan data for immunosuppressive conditions and hospitalizations for acute respiratory illness, United States. Emer Infect Dis 2020; 26(8):1720–30.

27. Tenforde MW, Patel MM, Ginde AA, et al. Effectiveness of Severe Acute Respiratory Syndrome Coronavirus 2 messenger RNA for preventing COVID-19 hospitalizations in the United States. Clin Infect Dis 2022; 74(9):1515–1524.

