## Supplementary Tables for "Effectiveness of COVID-19 Vaccines at Preventing Emergency Department or Urgent Care Encounters and Hospitalizations Among Immunocompromised Adults: An Observational Study of Real-World Data Across 10 US States from August-December 2021"

**Supplementary Table 1**. Immunocompromising conditions and related International Classification of Diseases, 9th and 10th Revision (ICD) discharge codes.

| Immunocompromising condition | ICD-10 codes | ICD-9 codes |
|---|---|---|
| **Solid malignancy** | | |
| Organ system malignant tumors | C00*-C80* | 140*-199* |
| Neuroendocrine tumors | C7A*, C7B*, D3A* | 209* |
| Encounter for antineoplastic radiation therapy | Z51.0 | V58.0* |
| Encounter for antineoplastic chemotherapy and immunotherapy | Z51.1* | V58.1* |
| **Hematological malignancy** | | |
| Hodgkin lymphoma | C81* | 201* |
| Follicular lymphoma | C82* | 202* |
| Non-follicular lymphoma | C83* | 200* |
| Malignant immunoproliferative diseases and certain other B-cell lymphomas | C88* | 203* |
| Multiple myeloma and malignant plasma cell neoplasms | C90* | 203* |
| Lymphoid leukemia | C91* | 204* |
| Myeloid leukemia | C92* | 205* |
| Monocytic leukemia | C93* | 206* |
| Other leukemias of specified cell type | C94* | 207* |
| Leukemia of unspecified cell type | C95* | 208* |
| Other and unspecified malignant neoplasms of lymphoid, hematopoietic and related tissue | C96* | 202* |
| Constitutional aplastic anemia | D61.0* | 284.0* |
| Idiopathic aplastic anemia | D61.2 | 284.89 |
| Aplastic anemia, unspecified | D61.9 | 284.9 |
| Congenital neutropenia | D70.0 | 288.01 |
| Functional disorders of polymorphuncular neutrophils | D71* | 288.1* |
| Mature T/NK-cell lymphoma | C84* | 202.1, 202.2, 202.7, 202.8, 200.6 |
| Other specified and unspecified types of non-Hodgkin lymphoma | C85* | 202.8* |
| Other specified types of T/NK cell lymphoma | C86* | 202.8* |
| Myelodysplastic syndromes | D46* | 238.72-238.75 |
| **Rheumatologic or inflammatory disorder** | | |
| Polyarteritis nodosa and related conditions | M30* | 446* |
| Lupus erythematosus | L93.0*, L93.2*, M32* | 695.4*, 710.0 |
| Diffuse connective tissue disease | L94*, M35.8*, M35.9* | 710, 710.5, 710.8, 710.9 |
| Rheumatoid arthritis with rheumatoid factor | M05* | |
| Other rheumatoid arthritis | M06* | |
| Juvenile arthritis | M08* | 714.3* |
| Sarcoidosis | D86* | 135* |
| Amyloidosis NOS | E85 | 277.3, 277.30 |
| Amyloidosis NEC | E85.1, E85.3, E85.8* | 277.39 |
| Multiple sclerosis | G35* | 340* |
| Allergic alveolitis/pneumonitis NOS | T78.40*, J67.9* | 495.9* |

| | | |
|---|---|---|
| Inflammatory spondylopathies | M46* | 720* |
| Polymyalgia rheumatic | M31.5*, M35.3* | 725* |
| Enteropathic arthropathies | M07* | |
| Wegener's granulomatosis | M31.3* | 446.4 |
| Dermatopolymyositis | M33* | 710.3, 710.4, |
| Systemic sclerosis (scleroderma) | M34* | 710.1 |
| Psoriatic juvenile arthropathy | L40.54 | 696.0* |
| Other psoriatic arthropathy | L40.59 | 696.0* |
| Heredofamilial amyloidosis, unspecified | E85.2 | 277.39 |
| Secondary systemic amyloidosis | E85.4 | 277.39 |
| Amyloidosis, unspecified | E85.9 | 277.30 |
| **Other intrinsic immune condition or immunodeficiency** | | |
| Disorders of immune mechanism | D89 | 279* |
| Other specified disorders involve the immune mechanism, not elsewhere classified | D89.8* | 279.49, 279.8 |
| Disorder involve the immune mechanism, unspecified | D89.9 | 279.9 |
| Leukocyte disease NEC | D72.89 | 288.8 |
| Leukocyte disease NOS | D27.9 | 288.9 |
| Immunodeficiency with predominantly antibody defects | D80* | 279.19 |
| Combined immunodeficiencies | D81 | 279.2, 279.13 |
| Severe combined immunodeficiency (SCID) with reticular dysgenesis | D81.0 | 279.2 |
| SCID with low T-and B-cell numbers | D81.1 | 279.2 |
| SCID with low or normal T-and B-cell numbers | D81.2 | 279.2 |
| Nezelof's syndrome | D81.4 | 279.13 |
| Other combined immunodeficiencies | D81.8* | 279.2 |
| Combined immunodeficiency, unspecified | D81.9 | 272.9 |
| Immunodeficiency associated with other major defects | D82* | 279.12, D79.11 |
| Common variable immunodeficiency | D83* | 279.06, 279.10 |
| Other immunodeficiencies | D84* | 277.6, 279* |
| Other constitutional aplastic anemia | D61.09 | 284.09 |
| End stage liver disease or equivalent (e.g., cirrhosis, ascities) | K70.3*, K70.4*, K72*, K74.3-K74.6, R18* | 571.2, 571.3, 571.5, 571.6, 572*, 789.5* |
| Nephrotic syndrome | N04* | 581* |
| Purine nucleoside phosphorylase [PNP] deficiency | D81.5 | |
| Major histocompatibility complex class I deficiency | D81.6 | 279.2 |
| Major histocompatibility complex class II deficiency | D81.7 | 279.2 |
| Polyclonal hypergammaglobulinemia | D89.0 | |
| Cryoglobulinemia | D89.1 | |
| Immune reconstitution syndrome | D89.3 | 279.8 |
| Mast cell activation syndrome and related disorders | D89.4* | 279.8 |
| **Organ or stem cell transplant** | | |
| Complications of bone marrow transplant | T86.0 | 996.85 |
| Complications of kidney transplant | T86.1 | |
| Complications of heart transplant | T86.2 | |
| Complications of heart-lung transplant | T86.3 | |
| Complications of liver transplant | T86.4 | |

| | | |
|---|---|---|
| Complications of stem cell transplant | T86.5 | 996.88 |
| Complications of lung transplant | T86.81 | |
| Complications of intestine transplant | T86.85 | |
| Transplanted organ and tissue status | Z94* | V42* |
| Transplanted organ removal status | Z98.85 | V45.87 |
| Post-transplant lymphoproliferative disorder | D47.Z1 | 238.77 |
| Encounter for aftercare following organ transplant | Z48.2* | V58.44 |

*** Includes all sub-codes.**

**Supplementary Table 2**. Covariates with remaining imbalances between vaccinated and unvaccinated groups after application of inverse propensity-to-be-vaccinated weighting for each vaccine effectiveness estimate, based on an absolute standardized mean or proportion difference ≥0.2.

| Vaccinated Exposure Group | Setting | Immunocompromised Status | Other Subgroup Criteria | Covariates with absolute SMD ≥0.2 after weighting* |
|---|---|---|---|---|
| 2 mRNA doses (≥14 days earlier) | ED/UC | Non-immunocompromised | None | None |
| 2 mRNA doses (14-149 days earlier) | ED/UC | Non-immunocompromised | None | None |
| 2 mRNA doses (≥150 days earlier) | ED/UC | Non-immunocompromised | None | None |
| 3 mRNA doses (≥7 days earlier) | ED/UC | Non-immunocompromised | None | None |
| 2 Moderna doses (≥14 days earlier) | ED/UC | Non-immunocompromised | None | None |
| 2 Moderna doses (14-149 days earlier) | ED/UC | Non-immunocompromised | None | None |
| 2 Moderna doses (≥150 days earlier) | ED/UC | Non-immunocompromised | None | None |
| 3 Moderna doses (≥7 days earlier) | ED/UC | Non-immunocompromised | None | None |
| 2 Pfizer doses (≥14 days earlier) | ED/UC | Non-immunocompromised | None | None |
| 2 Pfizer doses (14-149 days earlier) | ED/UC | Non-immunocompromised | None | None |
| 2 Pfizer doses (≥150 days earlier) | ED/UC | Non-immunocompromised | None | None |
| 3 Pfizer doses (≥7 days earlier) | ED/UC | Non-immunocompromised | None | None |
| 2 mRNA doses (≥14 days earlier) | ED/UC | Immunocompromised | None | None |
| 2 mRNA doses (14-149 days earlier) | ED/UC | Immunocompromised | None | None |
| 2 mRNA doses (≥150 days earlier) | ED/UC | Immunocompromised | None | None |
| 3 mRNA doses (≥7 days earlier) | ED/UC | Immunocompromised | None | None |
| 2 Moderna doses (≥14 days earlier) | ED/UC | Immunocompromised | None | None |
| 2 Moderna doses (14-149 days earlier) | ED/UC | Immunocompromised | None | None |
| 2 Moderna doses (≥150 days earlier) | ED/UC | Immunocompromised | None | None |
| 3 Moderna doses (≥7 days earlier) | ED/UC | Immunocompromised | None | Urban-rural classification of facility (SMD=0.20) |
| 2 Pfizer doses (≥14 days earlier) | ED/UC | Immunocompromised | None | None |
| 2 Pfizer doses (14-149 days earlier) | ED/UC | Immunocompromised | None | None |
| 2 Pfizer doses (≥150 days earlier) | ED/UC | Immunocompromised | None | None |
| 3 Pfizer doses (≥7 days earlier) | ED/UC | Immunocompromised | None | None |
| 2 mRNA doses (≥14 days earlier) | Hospital | Non-immunocompromised | None | None |
| 2 mRNA doses (14-149 days earlier) | Hospital | Non-immunocompromised | None | None |
| 2 mRNA doses (≥150 days earlier) | Hospital | Non-immunocompromised | None | None |
| 3 mRNA doses (≥7 days earlier) | Hospital | Non-immunocompromised | None | None |
| 2 Moderna doses (≥14 days earlier) | Hospital | Non-immunocompromised | None | None |
| 2 Moderna doses (14-149 days earlier) | Hospital | Non-immunocompromised | None | None |
| 2 Moderna doses (≥150 days earlier) | Hospital | Non-immunocompromised | None | None |
| 3 Moderna doses (≥7 days earlier) | Hospital | Non-immunocompromised | None | None |

| Vaccinated Exposure Group | Setting | Immunocompromised Status | Other Subgroup Criteria | Covariates with absolute SMD ≥0.2 after weighting* |
|---|---|---|---|---|
| 2 Pfizer doses (≥14 days earlier) | Hospital | Non-immunocompromised | None | None |
| 2 Pfizer doses (14-149 days earlier) | Hospital | Non-immunocompromised | None | None |
| 2 Pfizer doses (≥150 days earlier) | Hospital | Non-immunocompromised | None | None |
| 3 Pfizer doses (≥7 days earlier) | Hospital | Non-immunocompromised | None | None |
| 2 mRNA doses (≥14 days earlier) | Hospital | Immunocompromised | None | None |
| 2 mRNA doses (14-149 days earlier) | Hospital | Immunocompromised | None | None |
| 2 mRNA doses (≥150 days earlier) | Hospital | Immunocompromised | None | None |
| 3 mRNA doses (≥7 days earlier) | Hospital | Immunocompromised | None | None |
| 2 Moderna doses (≥14 days earlier) | Hospital | Immunocompromised | None | None |
| 2 Moderna doses (14-149 days earlier) | Hospital | Immunocompromised | None | None |
| 2 Moderna doses (≥150 days earlier) | Hospital | Immunocompromised | None | None |
| 3 Moderna doses (≥7 days earlier) | Hospital | Immunocompromised | None | None |
| 2 Pfizer doses (≥14 days earlier) | Hospital | Immunocompromised | None | None |
| 2 Pfizer doses (14-149 days earlier) | Hospital | Immunocompromised | None | None |
| 2 Pfizer doses (≥150 days earlier) | Hospital | Immunocompromised | None | None |
| 3 Pfizer doses (≥7 days earlier) | Hospital | Immunocompromised | None | None |
| 2 mRNA doses (≥14 days earlier) | ED/UC | Solid malignancy only | None | None |
| 3 mRNA doses (≥7 days earlier) | ED/UC | Solid malignancy only | None | None |
| 2 mRNA doses (≥14 days earlier) | ED/UC | Hematological malignancy only | None | None |
| 3 mRNA doses (≥7 days earlier) | ED/UC | Hematological malignancy only | None | None |
| 2 mRNA doses (≥14 days earlier) | ED/UC | Rheumatologic/inflammatory only | None | None |
| 3 mRNA doses (≥7 days earlier) | ED/UC | Rheumatologic/inflammatory only | None | None |
| 2 mRNA doses (≥14 days earlier) | ED/UC | Other immune disorder only | None | None |
| 3 mRNA doses (≥7 days earlier) | ED/UC | Other immune disorder only | None | Presence of prior SARS-CoV-2 testing pre-vaccination (SMD=0.27) |
| 2 mRNA doses (≥14 days earlier) | ED/UC | Organ/stem cell transplant only | None | Presence of prior SARS-CoV-2 testing pre-vaccination (SMD=0.21) |
| 3 mRNA doses (≥7 days earlier) | ED/UC | Organ/stem cell transplant only | None | Race (SMD=0.24) |
| 2 mRNA doses (≥14 days earlier) | Hospital | Solid malignancy only | None | None |
| 3 mRNA doses (≥7 days earlier) | Hospital | Solid malignancy only | None | None |
| 2 mRNA doses (≥14 days earlier) | Hospital | Hematological malignancy only | None | None |
| 3 mRNA doses (≥7 days earlier) | Hospital | Hematological malignancy only | None | Presence of prior SARS-CoV-2 testing pre-vaccination (SMD=0.29), ischemic heart disease (SMD=0.24) |
| 2 mRNA doses (≥14 days earlier) | Hospital | Rheumatologic/inflammatory only | None | None |
| 3 mRNA doses (≥7 days earlier) | Hospital | Rheumatologic/inflammatory only | None | None |

| Vaccinated Exposure Group | Setting | Immunocompromised Status | Other Subgroup Criteria | Covariates with absolute SMD ≥0.2 after weighting* |
|---|---|---|---|---|
| 2 mRNA doses (≥14 days earlier) | Hospital | Other immune disorder only | None | None |
| 3 mRNA doses (≥7 days earlier) | Hospital | Other immune disorder only | None | Hypertension (SMD=0.22) |
| 2 mRNA doses (≥14 days earlier) | Hospital | Organ/stem cell transplant only | None | Presence of prior SARS-CoV-2 testing pre-vaccination (SMD=0.23) |
| 3 mRNA doses (≥7 days earlier) | Hospital | Organ/stem cell transplant only | None | Other neurological/musculoskeletal disorder (SMD=0.27) |
| 2 mRNA doses (≥14 days earlier) | ED/UC | Immunocompromised, no organ/stem cell transplant | None | None |
| 3 mRNA doses (≥7 days earlier) | ED/UC | Immunocompromised, no organ/stem cell transplant | None | None |
| 2 Moderna doses (≥14 days earlier) | ED/UC | Immunocompromised, no organ/stem cell transplant | None | None |
| 3 Moderna doses (≥7 days earlier) | ED/UC | Immunocompromised, no organ/stem cell transplant | None | None |
| 2 Pfizer doses (≥14 days earlier) | ED/UC | Immunocompromised, no organ/stem cell transplant | None | None |
| 3 Pfizer doses (≥7 days earlier) | ED/UC | Immunocompromised, no organ/stem cell transplant | None | None |
| 2 mRNA doses (≥14 days earlier) | ED/UC | Organ/stem cell transplant | None | None |
| 3 mRNA doses (≥7 days earlier) | ED/UC | Organ/stem cell transplant | None | None |
| 2 Moderna doses (≥14 days earlier) | ED/UC | Organ/stem cell transplant | None | None |
| 3 Moderna doses (≥7 days earlier) | ED/UC | Organ/stem cell transplant | None | Other neurological/musculoskeletal disorder (SMD=0.20), COPD (SMD=0.22) |
| 2 Pfizer doses (≥14 days earlier) | ED/UC | Organ/stem cell transplant | None | None |
| 3 Pfizer doses (≥7 days earlier) | ED/UC | Organ/stem cell transplant | None | None |
| 2 mRNA doses (≥14 days earlier) | Hospital | Immunocompromised, no organ/stem cell transplant | None | None |
| 3 mRNA doses (≥7 days earlier) | Hospital | Immunocompromised, no organ/stem cell transplant | None | None |
| 2 Moderna doses (≥14 days earlier) | Hospital | Immunocompromised, no organ/stem cell transplant | None | None |
| 3 Moderna doses (≥7 days earlier) | Hospital | Immunocompromised, no organ/stem cell transplant | None | None |
| 2 Pfizer doses (≥14 days earlier) | Hospital | Immunocompromised, no organ/stem cell transplant | None | None |

| **Vaccinated Exposure Group** | **Setting** | **Immunocompromised Status** | **Other Subgroup Criteria** | **Covariates with absolute SMD ≥0.2 after weighting*** |
|---|---|---|---|---|
| 3 Pfizer doses (≥7 days earlier) | Hospital | Immunocompromised, no organ/stem cell transplant | None | None |
| 2 mRNA doses (≥14 days earlier) | Hospital | Organ/stem cell transplant | None | None |
| 3 mRNA doses (≥7 days earlier) | Hospital | Organ/stem cell transplant | None | None |
| 2 Moderna doses (≥14 days earlier) | Hospital | Organ/stem cell transplant | None | None |
| 3 Moderna doses (≥7 days earlier) | Hospital | Organ/stem cell transplant | None | Blood disorder (SMD=0.21), other heart disease (SMD=0.21), asthma (SMD=0.24) |
| 2 Pfizer doses (≥14 days earlier) | Hospital | Organ/stem cell transplant | None | None |
| 3 Pfizer doses (≥7 days earlier) | Hospital | Organ/stem cell transplant | None | Other metabolic disease (SMD=0.25) |
| 2 mRNA doses (≥14 days earlier) | ED/UC | Non-immunocompromised | Ages 18-64 | None |
| 3 mRNA doses (≥7 days earlier) | ED/UC | Non-immunocompromised | Ages 18-64 | None |
| 2 mRNA doses (≥14 days earlier) | ED/UC | Non-immunocompromised | Ages ≥65 | None |
| 3 mRNA doses (≥7 days earlier) | ED/UC | Non-immunocompromised | Ages ≥65 | None |
| 2 mRNA doses (≥14 days earlier) | ED/UC | Immunocompromised | Ages 18-64 | None |
| 3 mRNA doses (≥7 days earlier) | ED/UC | Immunocompromised | Ages 18-64 | Liver disease (SMD=0.21) |
| 2 mRNA doses (≥14 days earlier) | ED/UC | Immunocompromised | Ages ≥65 | None |
| 3 mRNA doses (≥7 days earlier) | ED/UC | Immunocompromised | Ages ≥65 | None |
| 2 mRNA doses (≥14 days earlier) | Hospital | Non-immunocompromised | Ages 18-64 | None |
| 3 mRNA doses (≥7 days earlier) | Hospital | Non-immunocompromised | Ages 18-64 | None |
| 2 mRNA doses (≥14 days earlier) | Hospital | Non-immunocompromised | Ages ≥65 | None |
| 3 mRNA doses (≥7 days earlier) | Hospital | Non-immunocompromised | Ages ≥65 | None |
| 2 mRNA doses (≥14 days earlier) | Hospital | Immunocompromised | Ages 18-64 | None |
| 3 mRNA doses (≥7 days earlier) | Hospital | Immunocompromised | Ages 18-64 | None |
| 2 mRNA doses (≥14 days earlier) | Hospital | Immunocompromised | Ages ≥65 | None |
| 3 mRNA doses (≥7 days earlier) | Hospital | Immunocompromised | Ages ≥65 | None |
| 2 mRNA doses (≥14 days earlier) | ED/UC | Non-immunocompromised | NH White | None |
| 3 mRNA doses (≥7 days earlier) | ED/UC | Non-immunocompromised | NH White | None |
| 2 mRNA doses (≥14 days earlier) | ED/UC | Non-immunocompromised | NH Black | None |
| 3 mRNA doses (≥7 days earlier) | ED/UC | Non-immunocompromised | NH Black | Chronic non-respiratory condition (SMD=0.25) |
| 2 mRNA doses (≥14 days earlier) | ED/UC | Non-immunocompromised | Hispanic | None |
| 3 mRNA doses (≥7 days earlier) | ED/UC | Non-immunocompromised | Hispanic | None |
| 2 mRNA doses (≥14 days earlier) | ED/UC | Immunocompromised | NH White | None |
| 3 mRNA doses (≥7 days earlier) | ED/UC | Immunocompromised | NH White | None |
| 2 mRNA doses (≥14 days earlier) | ED/UC | Immunocompromised | NH Black | None |
| 3 mRNA doses (≥7 days earlier) | ED/UC | Immunocompromised | NH Black | Liver disease (SMD=0.21), ischemic heart disease (SMD=0.22) |

| Vaccinated Exposure Group | Setting | Immunocompromised Status | Other Subgroup Criteria | Covariates with absolute SMD ≥0.2 after weighting* |
|---|---|---|---|---|
| 2 mRNA doses (≥14 days earlier) | ED/UC | Immunocompromised | Hispanic | None |
| 3 mRNA doses (≥7 days earlier) | ED/UC | Immunocompromised | Hispanic | None |
| 2 mRNA doses (≥14 days earlier) | Hospital | Non-immunocompromised | NH White | None |
| 3 mRNA doses (≥7 days earlier) | Hospital | Non-immunocompromised | NH White | None |
| 2 mRNA doses (≥14 days earlier) | Hospital | Non-immunocompromised | NH Black | None |
| 3 mRNA doses (≥7 days earlier) | Hospital | Non-immunocompromised | NH Black | None |
| 2 mRNA doses (≥14 days earlier) | Hospital | Non-immunocompromised | Hispanic | None |
| 3 mRNA doses (≥7 days earlier) | Hospital | Non-immunocompromised | Hispanic | Renal disease (SMD=0.25), clinical obesity (SMD=0.21), hypertension (SMD=0.26), other metabolic disease (SMD=0.31) |
| 2 mRNA doses (≥14 days earlier) | Hospital | Immunocompromised | NH White | None |
| 3 mRNA doses (≥7 days earlier) | Hospital | Immunocompromised | NH White | None |
| 2 mRNA doses (≥14 days earlier) | Hospital | Immunocompromised | NH Black | None |
| 3 mRNA doses (≥7 days earlier) | Hospital | Immunocompromised | NH Black | None |
| 2 mRNA doses (≥14 days earlier) | Hospital | Immunocompromised | Hispanic | None |
| 3 mRNA doses (≥7 days earlier) | Hospital | Immunocompromised | Hispanic | Hypertension (SMD=0.21) |
| 1 Janssen dose (≥14 days earlier) | ED/UC | Non-immunocompromised | None | None |
| 1 Janssen dose (14-149 days earlier) | ED/UC | Non-immunocompromised | None | None |
| 1 Janssen dose (≥150 days earlier) | ED/UC | Non-immunocompromised | None | None |
| 1 Janssen dose plus 2nd dose any product (≥7 days earlier) | ED/UC | Non-immunocompromised | None | Presence of prior SARS-CoV-2 testing pre-vaccination (SMD=0.22) |
| 1 Janssen dose (≥14 days earlier) | ED/UC | Immunocompromised | None | None |
| 1 Janssen dose (14-149 days earlier) | ED/UC | Immunocompromised | None | None |
| 1 Janssen dose (≥150 days earlier) | ED/UC | Immunocompromised | None | None |
| 1 Janssen dose plus 2nd dose any product (≥7 days earlier) | ED/UC | Immunocompromised | None | Solid malignancy (SMD=0.27), Rheumatologic or inflammatory disorder (SMD=0.22) |
| 1 Janssen dose (≥14 days earlier) | Hospital | Non-immunocompromised | None | None |
| 1 Janssen dose (14-149 days earlier) | Hospital | Non-immunocompromised | None | None |
| 1 Janssen dose (≥150 days earlier) | Hospital | Non-immunocompromised | None | None |
| 1 Janssen dose plus 2nd dose any product (≥7 days earlier) | Hospital | Non-immunocompromised | None | Heart failure (SMD=0.38), non-respiratory underlying medical condition (SMD=0.36), clinical obesity (SMD=0.30) |
| 1 Janssen dose (≥14 days earlier) | Hospital | Immunocompromised | None | None |
| 1 Janssen dose (14-149 days earlier) | Hospital | Immunocompromised | None | None |
| 1 Janssen dose (≥150 days earlier) | Hospital | Immunocompromised | None | None |

| Vaccinated Exposure Group | Setting | Immunocompromised Status | Other Subgroup Criteria | Covariates with absolute SMD ≥0.2 after weighting* |
|---|---|---|---|---|
| 1 Janssen dose plus 2nd dose any product (≥7 days earlier) | Hospital | Immunocompromised | None | COPD (SMD=0.35), rheumatologic or inflammatory disorder (SMD=0.21), chronic respiratory condition (SMD=0.33), other metabolic disease (SMD=0.31) |

Abbreviations: ED, emergency department; NH, non-Hispanic; SMD, standardized mean or proportion difference; UC, urgent care.
* Covariates considered for inclusion in propensity score models and evaluated for imbalances after inverse propensity-to-be-vaccinated weighting, as relevant, included age, sex, race, ethnicity, Medicaid status, calendar date, geographic region, local SARS-CoV-2 circulation on the day of each medical visit, urban-rural classification of facility, hospital type, number of hospital beds, hospital ownership, chronic respiratory condition, chronic non-respiratory condition, asthma, COPD, other chronic lung disease, heart failure, ischemic heart disease, hypertension, other heart disease, stroke, other cerebrovascular disease, diabetes type 1, diabetes type 2, diabetes due to underlying conditions or other specified diabetes, other metabolic disease (excluding diabetes), clinical obesity, clinical underweight, renal disease, liver disease, blood disorder, dementia, other neurological/musculoskeletal disorder, Down syndrome, HIV/AIDS, solid malignancy, hematological malignancy, rheumatologic or inflammatory disorder, other intrinsic immune disorder or underlying immunodeficiency, organ or stem cell transplant, presence of prior SARS-CoV-2 testing (pre-vaccination, if vaccinated), and prior positive SARS-CoV-2 test result (pre-vaccination, if vaccinated). An absolute SMD ≥0.20 indicates a non-negligible difference in variable distributions between events for vaccinated versus unvaccinated patients. All covariates with SMD ≥0.20 after weighting were also included in the multivariable logistic regression model for the respective vaccine effectiveness estimate(s) to minimize residual confounding.

**Supplementary Table 3**. Covariates with remaining imbalances between 3-dose mRNA-vaccinated and 2-dose mRNA-vaccinated groups after application of inverse propensity-to-be-vaccinated weighting for each relative vaccine effectiveness estimate, based on an absolute standardized mean or proportion difference ≥0.2.

| 3-Dose Vaccinated Exposure Group | 2-Dose Vaccinated Referent Group | Setting | Immunocompromised Status | Covariates with absolute SMD ≥0.2 after weighting* |
|---|---|---|---|---|
| 3 mRNA doses (≥7 days earlier) | 2 mRNA doses (≥14 days earlier) | ED/UC | Non-immunocompromised | None |
| 3 Moderna doses (≥7 days earlier) | 2 Moderna doses (≥14 days earlier) | ED/UC | Non-immunocompromised | None |
| 3 Pfizer doses (≥7 days earlier) | 2 Pfizer doses (≥14 days earlier) | ED/UC | Non-immunocompromised | None |
| 3 mRNA doses (≥7 days earlier) | 2 mRNA doses (≥14 days earlier) | ED/UC | Immunocompromised | None |
| 3 Moderna doses (≥7 days earlier) | 2 Moderna doses (≥14 days earlier) | ED/UC | Immunocompromised | None |
| 3 Pfizer doses (≥7 days earlier) | 2 Pfizer doses (≥14 days earlier) | ED/UC | Immunocompromised | None |
| 3 mRNA doses (≥7 days earlier) | 2 mRNA doses (≥14 days earlier) | Hospital | Non-immunocompromised | None |
| 3 Moderna doses (≥7 days earlier) | 2 Moderna doses (≥14 days earlier) | Hospital | Non-immunocompromised | None |
| 3 Pfizer doses (≥7 days earlier) | 2 Pfizer doses (≥14 days earlier) | Hospital | Non-immunocompromised | None |
| 3 mRNA doses (≥7 days earlier) | 2 mRNA doses (≥14 days earlier) | Hospital | Immunocompromised | None |
| 3 Moderna doses (≥7 days earlier) | 2 Moderna doses (≥14 days earlier) | Hospital | Immunocompromised | None |
| 3 Pfizer doses (≥7 days earlier) | 2 Pfizer doses (≥14 days earlier) | Hospital | Immunocompromised | None |

Abbreviations: ED, emergency department; SMD, standardized mean or proportion difference; UC, urgent care.

* Covariates considered for inclusion in propensity score models and evaluated for imbalances after inverse propensity-to-be-vaccinated weighting, as relevant, included age, sex, race, ethnicity, Medicaid status, calendar date, geographic region, local SARS-CoV-2 circulation on the day of each medical visit, urban-rural classification of facility, hospital type, number of hospital beds, hospital ownership, chronic respiratory condition, chronic non-respiratory condition, asthma, COPD, other chronic lung disease, heart failure, ischemic heart disease, hypertension, other heart disease, stroke, other cerebrovascular disease, diabetes type 1, diabetes type 2, diabetes due to underlying conditions or other specified diabetes, other metabolic disease (excluding diabetes), clinical obesity, clinical underweight, renal disease, liver disease, blood disorder, dementia, other neurological/musculoskeletal disorder, Down syndrome, HIV/AIDS, solid malignancy, hematological malignancy, rheumatologic or inflammatory disorder, other intrinsic immune disorder or underlying immunodeficiency, organ or stem cell transplant, presence of prior SARS-CoV-2 testing (pre-vaccination, if vaccinated), and prior positive SARS-CoV-2 test result (pre-vaccination, if vaccinated). An absolute SMD ≥0.20 indicates a non-negligible difference in variable distributions between events for 3-dose mRNA-vaccinated versus 2-dose mRNA-vaccinated patients. All covariates with SMD ≥0.20 after weighting were also included in the multivariable logistic regression model for the respective relative vaccine effectiveness estimate(s) to minimize residual confounding.

**Supplementary Table 4.** mRNA COVID-19 vaccine effectiveness against laboratory-confirmed COVID-19–associated emergency department or urgent care events and hospitalizations among non-immunocompromised and immunocompromised adults, by age group and number of vaccine doses, August–December 2021.

| | Emergency department and urgent care events | | | | | | Hospitalizations | | | | | |
|---|---|---|---|---|---|---|---|---|---|---|---|---|
| | Total | No. SARS-CoV-2 negative (col %) | No. SARS-CoV-2 positive (col %) | Proportion SARS-CoV-2 positive (row %) | Unadjusted VE* % (95% CI) | Adjusted VE† % (95% CI) | Total | No. SARS-CoV-2 negative (col %) | No. SARS-CoV-2 positive (col %) | Proportion SARS-CoV-2 positive (row %) | Unadjusted VE* % (95% CI) | Adjusted VE† % (95% CI) |
| **Non-immunocompromised adults‡** | | | | | | | | | | | | |
| **Aged 18-64 years** | | | | | | | | | | | | |
| Unvaccinated (referent) | 79740 | 52071 (45.8) | 27669 (83.4) | 34.7 | – | – | 20621 | 11862 (47.8) | 8759 (92.1) | 42.5 | – | – |
| Any mRNA vaccine | | | | | | | | | | | | |
| 2 doses (≥14 days earlier) | 60631 | 55331 (48.6) | 5300 (16.0) | 8.7 | 82 (81-83) | 82 (82-83) | 12296 | 11568 (46.6) | 728 (7.7) | 5.9 | 91 (91-92) | 91 (91-92) |
| 3 doses (≥7 days earlier) | 6551 | 6340 (5.6) | 211 (0.6) | 3.2 | 94 (93-95) | 94 (92-95) | 1434 | 1410 (5.7) | 24 (0.3) | 1.7 | 98 (97-98) | 97 (95-98) |
| **Aged ≥65 years** | | | | | | | | | | | | |
| Unvaccinated (referent) | 14373 | 8727 (20.0) | 5646 (59.6) | 39.3 | – | – | 11785 | 7270 (24.3) | 4515 (68.7) | 38.3 | – | – |
| Any mRNA vaccine | | | | | | | | | | | | |
| 2 doses (≥14 days earlier) | 28915 | 25349 (58.0) | 3566 (37.7) | 12.3 | 78 (77-79) | 79 (78-80) | 19595 | 17659 (58.9) | 1936 (29.4) | 9.9 | 82 (81-83) | 81 (80-83) |
| 3 doses (≥7 days earlier) | 9861 | 9607 (22.0) | 254 (2.7) | 2.6 | 96 (95-96) | 94 (93-95) | 5151 | 5027 (16.8) | 124 (1.9) | 2.4 | 96 (95-97) | 95 (93-96) |
| **Immunocompromised adults‡** | | | | | | | | | | | | |
| **Aged 18-64 years** | | | | | | | | | | | | |
| Unvaccinated (referent) | 1903 | 1482 (42.7) | 421 (69.2) | 22.1 | – | – | 3290 | 2568 (39.4) | 722 (72.5) | 21.9 | – | – |
| Any mRNA vaccine | | | | | | | | | | | | |
| 2 doses (≥14 days earlier) | 1710 | 1556 (44.8) | 154 (25.3) | 9.0 | 65 (58-71) | 65 (57-72) | 3389 | 3159 (48.4) | 230 (23.1) | 6.8 | 74 (70-78) | 73 (68-77) |
| 3 doses (≥7 days earlier) | 468 | 435 (12.5) | 33 (5.4) | 7.0 | 73 (61-82) | 68 (51-79) | 841 | 797 (12.2) | 44 (4.4) | 5.2 | 80 (73-86) | 69 (54-79) |
| **Aged ≥65 years** | | | | | | | | | | | | |
| Unvaccinated (referent) | 1295 | 1032 (24.4) | 263 (49.6) | 20.3 | – | – | 2542 | 2006 (19.6) | 536 (49.4) | 21.1 | – | – |
| Any mRNA vaccine | | | | | | | | | | | | |
| 2 doses (≥14 days earlier) | 2513 | 2284 (53.9) | 229 (43.2) | 9.1 | 61 (52-68) | 69 (62-75) | 6246 | 5771 (56.4) | 475 (43.8) | 7.6 | 69 (65-73) | 74 (69-77) |
| 3 doses (≥7 days earlier) | 959 | 921 (21.7) | 38 (7.2) | 4.0 | 84 (77-89) | 80 (70-87) | 2535 | 2461 (24.0) | 74 (6.8) | 2.9 | 89 (86-91) | 89 (84-92) |

Abbreviations: CI, confidence interval; VE, vaccine effectiveness.

*Unadjusted vaccine effectiveness was calculated as [1−odds ratio] x 100%, estimated using a test-negative design.
† Adjusted vaccine effectiveness was calculated as [1−odds ratio] x 100%, estimated using a test-negative design and adjusted for age, geographic region, calendar time, prior positive SARS-CoV-2 test result, and local SARS-CoV-2 circulation on the day of each medical visit, and weighted for patients' inverse propensity to be vaccinated (if the patient was vaccinated) or unvaccinated (if the patient was not vaccinated). Generalized boosted regression trees were used to estimate the propensity to be vaccinated based on sociodemographic characteristics, underlying medical conditions, known prior SARS-CoV-2 infection, and facility or hospital characteristics.
‡ Immunocompromised status was defined by the presence (presumed immunocompromised) or absence (presumed non-immunocompromised) of at least one discharge code from the International Classification of Diseases, 9th and 10th Revisions, for solid malignancy, hematologic malignancy, rheumatologic or inflammatory disorder, other intrinsic immune condition or immunodeficiency, or organ or stem cell transplant.

**Supplementary Table 5.** mRNA COVID-19 vaccine effectiveness against laboratory-confirmed COVID-19–associated emergency department or urgent care events and hospitalizations among non-immunocompromised and immunocompromised adults, by race/ethnicity and number of vaccine doses, August–December 2021.

| | Emergency department and urgent care events | | | | | | Hospitalizations | | | | | |
|---|---|---|---|---|---|---|---|---|---|---|---|---|
| | Total | No. SARS-CoV-2 negative (col %) | No. SARS-CoV-2 positive (col %) | Proportion SARS-CoV-2 positive (row %) | Unadjusted VE* % (95% CI) | VE† % (95% CI) | Total | No. SARS-CoV-2 negative (col %) | No. SARS-CoV-2 positive (col %) | Proportion SARS-CoV-2 positive (row %) | Unadjusted VE* % (95% CI) | VE† % (95% CI) |
| **Non-immunocompromised adults‡** | | | | | | | | | | | | |
| **Non-Hispanic white** | | | | | | | | | | | | |
| Unvaccinated (referent) | 57491 | 36896 (36.4) | 20595 (76.8) | 35.8 | – | – | 19882 | 11592 (32.9) | 8290 (80.4) | 41.7 | – | – |
| Any mRNA vaccine | | | | | | | | | | | | |
| 2 doses (≥14 days earlier) | 58706 | 52799 (52.2) | 5907 (22.0) | 10.1 | 80 (79-81) | 81 (80-82) | 20903 | 18992 (53.8) | 1911 (18.5) | 9.1 | 86 (85-87) | 85 (84-86) |
| 3 doses (≥7 days earlier) | 11834 | 11512 (11.4) | 322 (1.2) | 2.7 | 95 (94-96) | 94 (93-95) | 4792 | 4687 (13.3) | 105 (1.0) | 2.2 | 97 (96-97) | 96 (95-97) |
| **Non-Hispanic Black** | | | | | | | | | | | | |
| Unvaccinated (referent) | 10883 | 7569 (52.9) | 3314 (84.8) | 30.4 | | – | 4394 | 3035 (47.5) | 1359 (87.3) | 30.9 | – | – |
| Any mRNA vaccine | | | | | | | | | | | | |
| 2 doses (≥14 days earlier) | 6526 | 5955 (41.6) | 571 (14.6) | 8.8 | 78 (76-80) | 78 (76-80) | 3137 | 2944 (46.1) | 193 (12.4) | 6.2 | 85 (83-88) | 86 (83-88) |
| 3 doses (≥7 days earlier) | 813 | 792 (5.5) | 21 (0.5) | 2.6 | 94 (91-96) | 90 (84-94) | 413 | 408 (6.4) | 5 (0.3) | 1.2 | 97 (93-99) | 94 (83-98) |
| **Hispanic** | | | | | | | | | | | | |
| Unvaccinated (referent) | 14865 | 9804 (41.9) | 5061 (81.8) | 34.0 | – | – | 4407 | 2509 (38.2) | 1898 (28.9) | 43.1 | – | – |
| Any mRNA vaccine | | | | | | | | | | | | |
| 2 doses (≥14 days earlier) | 13101 | 12002 (51.3) | 1099 (17.8) | 8.4 | 82 (81-83) | 84 (83-86) | 3818 | 3563 (54.2) | 255 (3.9) | 6.7 | 91 (89-92) | 92 (90-93) |
| 3 doses (≥7 days earlier) | 1632 | 1603 (6.8) | 29 (0.5) | 1.8 | 96 (95-98) | 96 (93-97) | 512 | 504 (7.7) | 8 (0.1) | 1.6 | 98 (96-99) | 96 (91-98) |
| **Immunocompromised adults‡** | | | | | | | | | | | | |
| **Non-Hispanic white** | | | | | | | | | | | | |
| Unvaccinated (referent) | 2045 | 1579 (30.6) | 466 (58.4) | 22.8 | – | – | 3737 | 2897 (26.2) | 840 (59.2) | 22.5 | – | – |
| Any mRNA vaccine | | | | | | | | | | | | |
| 2 doses (≥14 days earlier) | 2871 | 2596 (50.3) | 275 (34.5) | 9.6 | 64 (58-69) | 68 (61-73) | 6370 | 5876 (53.1) | 494 (34.8) | 7.8 | 71 (67-74) | 72 (68-75) |

| | | | | | | | | | | | | |
|---|---|---|---|---|---|---|---|---|---|---|---|---|
| 3 doses (≥7 days earlier) | 1041 | 984 (19.1) | 57 (7.1) | 5.5 | 80 (74-85) | 70 (57-79) | 2374 | 2288 20.7) | 86 (6.1) | 3.6 | 87 (84-90) | 82 (76-87) |
| **Non-Hispanic Black** | | | | | | | | | | | | |
| Unvaccinated (referent) | 463 | 396 (43.0) | 67 (68.4) | 14.5 | – | – | 760 | 625 (34.6) | 135 (63.7) | 17.8 | – | – |
| Any mRNA vaccine | | | | | | | | | | | | |
| 2 doses (≥14 days earlier) | 452 | 426 (46.3) | 26 (26.5) | 5.8 | 64 (42-78) | 66 (41-80) | 1013 | 948 (52.5) | 65 (30.7) | 6.4 | 68 (57-77) | 73 (62-82) |
| 3 doses (≥7 days earlier) | 104 | 99 (10.7) | 5 (5.1) | 4.8 | 70 (24-88) | 55 (-65-88) | 245 | 233 (12.9) | 12 (5.7) | 4.9 | 76 (56-87) | 82 (56-92) |
| **Hispanic** | | | | | | | | | | | | |
| Unvaccinated (referent) | 456 | 356 (35.5) | 100 (63.7) | 21.9 | – | – | 800 | 637 (29.4) | 163 (59.5) | 20.4 | – | – |
| Any mRNA vaccine | | | | | | | | | | | | |
| 2 doses (≥14 days earlier) | 551 | 501 (49.9) | 50 (31.8) | 9.1 | 64 (49-75) | 69 (51-81) | 1252 | 1156 (53.4) | 96 (35.0) | 7.7 | 68 (57-75) | 70 (58-78) |
| 3 doses (≥7 days earlier) | 154 | 147 (14.6) | 7 (4.5) | 4.5 | 83 (63-92) | 87 (56-96) | 387 | 372 (17.2) | 15 (5.5) | 3.9 | 84 (73-91) | 82 (65-91) |

Abbreviations: CI, confidence interval; VE, vaccine effectiveness.

*Unadjusted vaccine effectiveness was calculated as [1−odds ratio] x 100%, estimated using a test-negative design.

† Adjusted vaccine effectiveness was calculated as [1−odds ratio] x 100%, estimated using a test-negative design and adjusted for age, geographic region, calendar time, prior positive SARS-CoV-2 test result, and local SARS-CoV-2 circulation on the day of each medical visit, and weighted for patients' inverse propensity to be vaccinated (if the patient was vaccinated) or unvaccinated (if the patient was not vaccinated). Generalized boosted regression trees were used to estimate the propensity to be vaccinated based on sociodemographic characteristics, underlying medical conditions, known prior SARS-CoV-2 infection, and facility or hospital characteristics.

‡Immunocompromised status was defined by the presence (presumed immunocompromised) or absence (presumed non-immunocompromised) of at least one discharge code from the International Classification of Diseases, 9th and 10th Revisions, for solid malignancy, hematologic malignancy, rheumatologic or inflammatory disorder, other intrinsic immune condition or immunodeficiency, or organ or stem cell transplant.

**Supplementary Table 6.** Characteristics of emergency department and urgent care events among non-immunocompromised and immunocompromised adults with COVID-19–like illness who received the Janssen (Ad26.COV2) COVID-19 vaccine as their first dose, VISION Network, 10 states, August–December 2021.

| | **Non-immunocompromised adults*** | **Immunocompromised adults*** |
|---|---|---|
| | **n (column %)** | **n (column %)** |
| **All ED/UC events** | 9,833 (100) | 496 (100) |
| **Site** | | |
| Baylor Scott & White Health | 1,034 (10.5) | 163 (32.9) |
| Columbia University† | 182 (1.8) | 31 (6.2) |
| HealthPartners† | 364 (3.7) | 9 (1.8) |
| Intermountain Healthcare | 3,139 (31.9) | 132 (26.6) |
| Kaiser Permanente Northern California | 2,179 (22.2) | 49 (9.9) |
| Kaiser Permanente Northwest | 913 (9.3) | 82 (16.5) |
| Regenstrief Institute | 1,443 (14.7) | 26 (5.2) |
| University of Colorado | 579 (5.9) | 4 (0.8) |
| **Age group, years** | | |
| 18-49 | 4,664 (47.4) | 101 (20.4) |
| 50-64 | 2,928 (29.8) | 184 (37.1) |
| 65-74 | 1,211 (12.3) | 132 (26.6) |
| 75-84 | 678 (6.9) | 60 (12.1) |
| ≥85 | 352 (3.6) | 19 (3.8) |
| **Sex** | | |
| Male | 4,291 (43.6) | 246 (49.6) |
| Female | 5,542 (56.4) | 250 (50.4) |
| **Race/ethnicity** | | |
| White, non-Hispanic | 6,523 (66.3) | 341 (68.7) |
| Black, non-Hispanic | 857 (8.7) | 63 (12.7) |
| Hispanic | 1,353 (13.8) | 58 (11.7) |
| Other, non-Hispanic‡ | 573 (5.8) | 19 (3.8) |
| Unknown | 527 (5.4) | 15 (3.0) |
| **Chronic respiratory condition§** | | |
| Yes | 1,936 (19.7) | 252 (50.8) |
| No | 7,897 (80.3) | 244 (49.2) |
| **Number of doses‖** | | |
| One Janssen dose | 9,179 (93.3) | 465 (93.7) |
| Two doses (any product as second dose) | 654 (6.6) | 31 (6.2) |
| mRNA vaccine as second dose | 440 (4.5) | 24 (4.8) |
| Janssen vaccine as second dose | 214 (2.2) | 7 (1.4) |

Abbreviations: ED, emergency department; UC, urgent care.

* Immunocompromised status at medical events was defined by the presence (presumed immunocompromised) or absence (presumed non-immunocompromised) of at least one discharge code from the International Classification of Diseases, 9th and 10th Revisions, for solid malignancy, hematologic malignancy, rheumatologic or inflammatory disorder, other intrinsic immune condition or immunodeficiency, or organ or stem cell transplant.

† ED data at Columbia University Irving Medical Center and HealthPartners exclude encounters that were transferred to an inpatient setting.

‡ Other race includes Asian, Native Hawaiian, or other Pacific islander, American Indian or Alaska Native, other not listed, and multiple races.

§ Chronic respiratory condition at discharge was defined as the presence of discharge codes from the International Classification of Diseases, 9th and 10th Revisions, for asthma, chronic obstructive pulmonary disease, or other lung disease.

|| Patients who received the Janssen vaccine as their first dose were classified according to receipt of the first in a 1-dose series ≥14 days before the medical event index date or the second dose (any product) in a 2-dose series ≥7 days before the index date. Patients who received two doses were further classified based on whether the second dose received was an mRNA vaccine or a second dose of the Janssen vaccine.

**Supplementary Table 7.** Characteristics of hospitalizations among non-immunocompromised and immunocompromised adults with COVID-19–like illness who received the Janssen (Ad26.COV2) COVID-19 vaccine as their first dose, VISION Network, 10 states, August–December 2021.

| | **Non-immunocompromised adults*** | **Immunocompromised adults*** |
|---|---|---|
| | **n (column %)** | **n (column %)** |
| **All hospitalizations** | 3,234 (100) | 968 (100) |
| **Site** | | |
| Baylor Scott & White Health | 518 (16.0) | 175 (18.1) |
| Columbia University | 138 (4.3) | 55 (5.7) |
| HealthPartners | 45 (1.4) | 18 (1.9) |
| Intermountain Healthcare | 334 (10.3) | 64 (6.6) |
| Kaiser Permanente Northern California | 968 (29.9) | 342 (35.3) |
| Kaiser Permanente Northwest | 191 (5.9) | 58 (6.0) |
| Regenstrief Institute | 727 (22.5) | 131 (13.5) |
| University of Colorado | 313 (9.7) | 125 (12.9) |
| **Age group, years** | | |
| 18-49 | 724 (22.4) | 142 (14.7) |
| 50-64 | 1076 (33.3) | 396 (40.9) |
| 65-74 | 665 (20.5) | 241 (24.9) |
| 75-84 | 464 (14.3) | 131 (13.5) |
| ≥85 | 305 (9.4) | 58 (6.0) |
| **Sex** | | |
| Male | 1555 (48.1) | 501 (51.8) |
| Female | 1679 (51.9) | 467 (48.2) |
| **Race/ethnicity** | | |
| White, non-Hispanic | 2046 (63.3) | 630 (65.0) |
| Black, non-Hispanic | 422 (13.0) | 114 (11.8) |
| Hispanic | 431 (13.3) | 131 (13.5) |
| Other, non-Hispanic† | 171 (5.3) | 63 (6.5) |
| Unknown | 164 (5.1) | 30 (3.1) |
| **Chronic respiratory condition‡** | | |
| Yes | 2106 (65.1) | 638 (65.9) |
| No | 1128 (34.9) | 330 (34.1) |
| **Number of doses§** | | |
| One Janssen dose | 3028 (93.6) | 920 (95.0) |
| Two doses (any product as second dose) | 206 (6.4) | 48 (5.0) |
| mRNA vaccine as second dose | 129 (4.0) | 33 (3.4) |
| Janssen vaccine as second dose | 77 (2.4) | 15 (1.5) |

* Immunocompromised status at hospitalizations was defined by the presence (presumed immunocompromised) or absence (presumed non-immunocompromised) of at least one discharge code from the International Classification of Diseases, 9th and 10th Revisions, for solid malignancy, hematologic malignancy, rheumatologic or inflammatory disorder, other intrinsic immune condition or immunodeficiency, or organ or stem cell transplant.

† Other race includes Asian, Native Hawaiian, or other Pacific islander, American Indian or Alaska Native, other not listed, and multiple races.

‡ Chronic respiratory condition at discharge was defined as the presence of discharge codes from the International Classification of Diseases, 9th and 10th Revisions, for asthma, chronic obstructive pulmonary disease, or other lung disease.

§ Patients who received the Janssen vaccine as their first dose were classified according to receipt of the first in a 1-dose series ≥14 days before the hospitalization index date or the second dose (any product) in a 2-dose series ≥7 days before the index date. Patients who received two doses were further classified based on whether the second dose received was an mRNA vaccine or a second dose of the Janssen vaccine.

**Supplementary Table 8.** Janssen (Ad26.COV2) COVID-19 vaccine effectiveness against laboratory-confirmed COVID-19–associated emergency department or urgent care events and hospitalizations among non-immunocompromised and immunocompromised adults, by number of vaccine doses, August–December 2021.

| | Emergency department or urgent care events | | | | | | Hospitalizations | | | | | |
|---|---|---|---|---|---|---|---|---|---|---|---|---|
| | **Total** | **No. SARS-CoV-2 negative (col %)** | **No. SARS-CoV-2 positive (col %)** | **Proportion SARS-CoV-2 positive (row %)** | **Unadjusted VE* % (95% CI)** | **Adjusted VE† % (95% CI)** | **Total** | **No. SARS-CoV-2 negative (col %)** | **No. SARS-CoV-2 positive (col %)** | **Proportion SARS-CoV-2 positive (row %)** | **Unadjusted VE* % (95% CI)** | **Adjusted VE† % (95% CI)** |
| **Non-immunocompromised‡** | | | | | | | | | | | | |
| Unvaccinated (ref) | 94113 | 60798 (87.9) | 33315 (95.7) | 35.4 | – | – | 32406 | 19132 (87.3) | 13274 (96.7) | 41.0 | – | – |
| 1 Janssen dose (≥14 days earlier) | 9179 | 7733 (11.2) | 1446 (4.2) | 15.8 | 66 (64-68) | 64 (61-66) | 3028 | 2580 (11.8) | 448 (3.3) | 14.8 | 75 (72-77) | 72 (69-76) |
| 1 Janssen dose (14-149 days earlier) | 2720 | 2297 (3.3) | 423 (1.2) | 15.6 | 66 (63-70) | 67 (63-71) | 945 | 824 (3.8) | 121 (0.9) | 12.8 | 79 (74-83) | 77 (71-82) |
| 1 Janssen dose (≥150 days earlier) | 6459 | 5436 (7.9) | 1023 (2.9) | 15.8 | 66 (63-68) | 62 (58-65) | 2083 | 1756 (8.0) | 327 (2.4) | 15.7 | 73 (70-76) | 69 (65-73) |
| 2 doses, 1 Janssen dose plus 2nd dose any product (≥7 days earlier) | 654 | 620 (0.9) | 34 (0.1) | 5.2 | 90 (86-93) | 90 (86-93) | 206 | 196 (0.9) | 10 (0.1) | 4.8 | 93 (86-96) | 93 (86-96) |
| **Immunocompromised‡** | | | | | | | | | | | | |
| Unvaccinated (ref) | 3198 | 2514 (85.6) | 684 (90.4) | 21.4 | – | – | 5832 | 4574 (84.0) | 1258 (92.7) | 21.6 | – | – |
| 1 Janssen dose (≥14 days earlier) | 465 | 395 (13.4) | 70 (9.2) | 15.1 | 35 (15-50) | 7 (-26-31) | 920 | 826 (15.2) | 94 (6.9) | 10.2 | 59 (48-67) | 45 (29-57) |
| 1 Janssen dose (14-149 days earlier) | 121 | 107 (3.6) | 14 (1.8) | 11.6 | 52 (16-73) | 9 (-66-50) | 238 | 211 (3.9) | 27 (2.0) | 11.3 | 53 (30-69) | 35 (-3-60) |
| 1 Janssen dose (≥150 days earlier) | 344 | 288 (9.8) | 56 (7.4) | 16.3 | 29 (4-47) | -6 (-49-25) | 682 | 615 (11.3) | 67 (4.9) | 9.8 | 60 (49-69) | 43 (23-58) |
| 2 doses, 1 Janssen dose plus 2nd dose any product (≥7 days earlier) | 31 | 28 (0.9) | 3 (0.4) | 9.7 | 61 (-30-88) | 71 (6-91) | 48 | 43 (0.8) | 5 (0.4) | 10.4 | 58 (-7-83) | 70 (25-88) |

Abbreviations: CI, confidence interval; VE, vaccine effectiveness.

*Unadjusted vaccine effectiveness was calculated as [1−odds ratio] x 100%, estimated using a test-negative design.

† Adjusted vaccine effectiveness was calculated as [1−odds ratio] x 100%, estimated using a test-negative design and adjusted for age, geographic region, calendar time, prior positive SARS-CoV-2 test result, and local SARS-CoV-2 circulation on the day of each medical visit, and weighted for patients' inverse propensity to be vaccinated (if the patient was vaccinated) or unvaccinated (if the patient was not vaccinated). Generalized boosted regression trees were used to estimate the propensity to be vaccinated based on sociodemographic characteristics, underlying medical conditions, known prior SARS-CoV-2 infection, and facility or hospital characteristics.

‡ Immunocompromised status was defined by the presence (presumed immunocompromised) or absence (presumed non-immunocompromised) of at least one discharge code from the International Classification of Diseases, 9th and 10th Revisions, for solid malignancy, hematologic malignancy, rheumatologic or inflammatory disorder, other intrinsic immune condition or immunodeficiency, or organ or stem cell transplant.
